# Development and Validation of Dementia Diagnosis in Adults through Machine Learning Frameworks: A Cross-Sectional Study Using Clinical and Imaging Data

**DOI:** 10.64898/2026.01.24.26344772

**Authors:** Fahad Mostafa, Kushagra Sharma, Hafiz Khan

## Abstract

**Background:** Dementia risk stratifications and diagnosis can significantly improve care planning and patient outcomes while delaying progression. Machine learning algorithms can identify patterns in clinical and neuroimaging data that may aid in the early-stage detection of dementia risk factors.

**Objective:** To evaluate the performance of the ensemble machine learning pipeline for classifying dementia status utilizing demographic, clinical, and imaging features, and to identify the most predictive variables contributing to model accuracy.

**Methods:** A cross-sectional study analyzed 373 MRI scans from 150 subjects aged 60–98 years. Variables included cognitive scores (MMSE, CDR), volumetric brain measures (eTIV, nWBV, ASF), demographic features (age, gender, education), and socioeconomic status. After preprocessing and imputing missing values with random forests, tree-based variable selection was performed, and the dataset was split into training and test sets, with 5-fold cross-validation used for model validation. An ensemble of 8 machine learning models was used to classify patients as demented or non-demented.

**Results:** Model performance was assessed using the area under the receiver operating characteristic (ROC) curve (AUC), accuracy, sensitivity, specificity, precision, F1 score, and Matthews Correlation Coefficient (MCC). Random Forest achieved the highest AUC (0.963), while MLP demonstrated the highest accuracy (94.6%), F1-score (0.943), and MCC (0.893). CDR, MMSE, and ASF were identified as the top predictors. Performance was robust across folds in 5-fold CV, and feature importance analyses supported clinical relevance.

**Conclusions:** Ensemble ML approaches offer high predictive performance in dementia classification. ML frameworks have the potential to be integrated into diagnostic support tools, enabling more accurate and earlier detection of dementia using clinical and imaging data.

## INTRODUCTION

According to the World Health Organization (WHO), over 46.8 million people have dementia around the world, and this number is predicted to grow to 74.7 million by 2030 and to 131.5 million by 2050 as the population ages [1]. Dementia is a syndrome primarily caused by Alzheimer’s disease (AD), accounting for 60-70% of dementia cases, especially in people over the age of 65 [2], and it affects the memory, thinking, learning capacity, comprehension, as well as the ability to perform everyday tasks. Dementia is a progressive condition that often begins at a stage known as Mild Cognitive Impairment (MCI), a period between normal aging and dementia during which individuals experience declines in cognitive abilities, primarily memory; they are still able to perform everyday tasks. Most individuals with MCI eventually progress to a complete dementia diagnosis. Research shows that early and accurate diagnosis of dementia can allow for personalized care, cost savings, better management of symptoms, and slow the progression of the syndrome [3].

Although no single exact cause for dementia has been identified, it is believed that a combination of a range of independent factors, such as age, genetics, health conditions, and lifestyle choices, is heavily linked to causing dementia. Dementia diagnosis involves comprehensive assessments like Mini-Mental State Examination (MMSE) along with structural brain changes like cerebral atrophy which are measured by Magnetic Resonance Imaging (MRI) scan [4]. Integrating these factors and assessments can be difficult for clinicians to make an early diagnosis, and traditional methods can be limited in their ability to capture the complex and multifactorial nature of the syndrome.

Using concurrent technologies, such as machine learning (ML), can help diagnose dementia by handling large datasets and developing predictive models. These models can identify patterns and interactions among variables and have shown promising results in improving speed, precision, and accuracy for early disease diagnosis across several fields, such as Parkinson’s and cardiovascular disease [5,6]. As more data are gathered, researchers develop ML techniques to build robust, accurate predictive models for dementia diagnosis.

In the last few years, several new machine learning techniques, such as supervised learning, deep learning, and artificial intelligence (AI), have been developed by researchers for the early-stage detection and diagnosis of dementia. Earlier this year, researchers at Birmingham City University developed deep learning methods to detect AD [7] accurately. In the study, the researchers used a framework that included separate trained models for structured data (clinical and cognitive information) and MRI. For structured data, the researchers used a hybrid model of Long Short-Term Memory (LSTM) and Feedforward Neural Network (FNN), while for MRI data, they used pre-trained deep learning methods: MobileNetV2 and ResNet50. The models were evaluated on two datasets: the National Alzheimer’s Coordinating Centre (NACC) and the Alzheimer’s Disease Neuroimaging Initiative (ADNI). The MRI-based model achieved 96.19% accuracy on the ADNI dataset, and the hybrid model obtained 99.82% accuracy. In a similar study, researchers at G.G. Vishwavidyalaya used hybrid deep learning algorithms to overcome challenges in the early detection of AD [8]. The researchers began by pre-processing images using Improved Adaptive Wiener Filtering (IAWF), a sophisticated noise-reduction technique applied to raw medical images such as MRIs. Next, they developed a hybrid feature-extraction method that combined Principal Component Analysis (PCA) with a Normalized Global Image Descriptor (NGIST). Next, the best features were selected using the Improved Wild Horse Optimization (IWHO) algorithm. Finally, the disease was diagnosed using a hybrid approach combining a Bi-directional Long Short-Term Memory (BiLSTM) network and a traditional Artificial Neural Network (ANN). The impressive accuracy reported for the models is 99.22% on the ADNI dataset and 98.96% on the Open Access Series of Imaging Studies (OASIS). Another study by the Department of Health, Blekinge Institute of Technology in Sweden, examined a statistical and ML-based hybrid system for predicting dementia [9]. The goal of the study was to develop a non-invasive, accurate system that could detect dementia early using patients’ health records. For the study, the authors used a large dataset from the Swedish National Study on Aging and Care (SNAC), which included 43,040 patient samples, each containing 75 features. The researchers used the F-score to rank features by their importance and selected the most descriptive ones. For classification, they developed an ensemble voting classifier based on five different ML models: Random Forest, Naive Bayes, Logistic Regression, Decision Tree, Support Vector Machines, and used a 5-fold cross-validation. The results from the hybrid system model achieved the highest accuracy of 98.25%, sensitivity of 97.44%, specificity of 95.74%, and MCC of 0.7535. Another study performed by the researchers at Seoul University used a two-layer model to help early diagnosis of dementia using machine learning techniques like Naive Bayes, Bayes Network, Begging, Logistic Regression (LR), Random Forest (RF), Support Vector Machine (SVM) and Multilayer Perceptron (MLP) and comparing the models using Precision, Recall, and F-measure [10]. Rossini et al. (2022) propose that combining machine learning with graph-theoretic analysis of integrated biomarkers, especially EEG signals, is a promising and cost-effective method for early dementia diagnosis. This innovative approach can identify individuals with Mild Cognitive Impairment (MCI) at high risk of progression, enabling personalized risk evaluation and facilitating timely interventions [11]. Nori et al. (2019) developed a machine learning model using a "label learning" approach on administrative and EHR data to correct for inaccurate diagnostic coding when predicting future dementia onset. Their model predicted incident dementia within 2 years with 47% sensitivity and an area under the curve of 87%, demonstrating its utility for screening patients for clinical trials and management [12]. Ahamed et al. (2020) developed a machine learning framework to identify the onset of dementia from IoT sensor data tracking daily life activities. By extracting and ranking features related to task duration and completion, their Fine Decision Tree model achieved 90.74% accuracy in distinguishing cognitively impaired individuals [13].

The early diagnosis of dementia, particularly AD, remains a critical challenge in neurodegenerative research. Functional magnetic resonance imaging (fMRI) has emerged as a valuable tool for capturing brain connectivity patterns that precede structural degeneration. Machine learning methods have been increasingly applied to analyze fMRI data to improve early diagnostic accuracy. SVMs and Convolutional Neural Networks (CNNs) have demonstrated promising results in classifying mild cognitive impairment (MCI) and early-stage AD from healthy controls. For example, Farheen Ramzan et al. (2020) integrated SVM with deep learning for feature representation, achieving high classification accuracy using resting-state fMRI data [14]. Similarly, Jinlong Hu et al. (2017) employed deep CNNs on 2D slices of fMRI volumes, significantly improving predictive performance [15]. Graph-based ML approaches, which model functional connectivity as networks, have shown potential in identifying subtle topological disruptions in brain regions. Thushara, A., Amma, C. U., & John, A. (2023) used graph theory-based metrics as features for ML classifiers, revealing characteristic patterns in MCI patients [16]. More recently, some researchers introduced graph convolutional networks (GCNs) to exploit inter-regional correlations, enhancing early dementia prediction. Another promising direction is the integration of multimodal data. Combining fMRI with structural MRI or genetic information through ensemble ML frameworks has been shown to improve robustness and generalizability. These hybrid models can better capture the complex neuropathology of dementia [17, 18, 19]. Despite advances, challenges persist, including data heterogeneity, small sample sizes, and the interpretability of ML models. Future research should focus on longitudinal data analysis and developing explainable AI models to translate ML findings into clinical applications.

In this study, we developed, compared, and validated interpretable machine learning–based ensemble frameworks using a cross-sectional dataset comprising demographic, cognitive, and neuroimaging variables associated with dementia disease risk which had not been looked at before. Key predictors included the Mini-Mental State Examination, Clinical Dementia Rating, age, gender, and socioeconomic status, as well as volumetric brain imaging measures. By integrating these complementary features, the models were designed to capture both functional and structural dimensions of disease progression. The overarching goal of this work was to establish a clinically relevant tool that assists in diagnosis and supports clinical decision-making by improving the accuracy and reliability of dementia classification.

## MATERIALS AND METHODS

### Data source and study population

The longitudinal dataset, comprising 373 observations and 15 variables, originates from a brain imaging study and is designed to identify potential factors associated with AD, the leading cause of dementia [30]. It includes demographic (age, gender), clinical (MMSE, CDR), and anatomical (eTIV, nWBV, ASF) measures, providing a comprehensive foundation for statistical analysis. Although the dataset contains multiple entries for individuals due to repeated clinical visits, robust analysis requires focusing on the most recent visit per subject to ensure data independence and reliability. To ensure data independence and avoid temporal bias, only the final visit per participant was used to train the model in a cross-sectional design. In this study, the longitudinal dataset was converted into a cross-sectional design by selecting only the final visit for each participant to ensure independent observations for model training. The cohort consists of 150 unique right-handed subjects aged 60 to 98 years, each with up to five visits, with MMSE scores ranging from 4 to 30 and CDR scores from 0 to 2, reflecting a broad spectrum of cognitive function and dementia severity (**Table 1**). Each participant underwent two or more study visits, separated by at least one year, resulting in a total of 373 imaging sessions, with three to four T1-weighted MRI scans acquired per visit in the clinic. The cohort includes both men and women and represents a broad spectrum of cognitive status. Of the 150 subjects, 72 remained consistently nondemented throughout the study, while 64 were diagnosed as demented at their initial visit and retained this diagnosis over time, including 51 individuals with mild to moderate Alzheimer’s disease. An additional 14 subjects initially classified as converted subsequently considered to dementia during follow-up, providing a valuable subset for studying disease progression and early structural biomarkers of dementia.

**Table 1.** Dementia coding variables.

| Variable Name | Variable Type | Variable Description | Range / Categories |
| --- | --- | --- | --- |
| Group | Categorical | Dementia group classification | Demented, Nondemented, Converted |
| Visit | Numeric | Visit number | 1 – 5 |
| MR Delay | Numeric | Days between baseline and MRI | 0 – 2639 |
| Gender | Categorical | Biological sex | M = Male, F = Female |
| Age | Numeric | Age in years | 60 – 98 |
| SES | Categorical | Socioeconomic status | 1 – 5 |
| EDUC | Categorical | Years of education | 6 – 23 |
| MMSE | Numeric | Mini-Mental State Examination score | 4 – 30 (normal $\geq 27$) |
| CDR | Numeric | Clinical Dementia Rating | 0 – 2 |
| eTIV | Numeric | Estimated Total Intracranial Volume (cc) | 1106 – 2004 |
| nWBV | Numeric | Normalized Whole Brain Volume | 0.644 – 0.837 |
| ASF | Numeric | Atlas Scaling Factor | 0.876 – 1.587 |
| Subject ID | Identifier | Unique patient ID | Alphanumeric |
| MRI ID | Identifier | Unique MRI scan identifier | Alphanumeric |
| Hand | Categorical | Handedness | All "R" (Right-handed) |

The estimated Total Intracranial Volume (eTIV) normalized Whole Brain Volume (nWBV), and Atlas Scaling Factor (ASF) were computed using automated volumetric analysis within the FreeSurfer processing pipeline, as part of the OASIS (Open Access Series of Imaging Studies) dataset. ASF represents the volumetric scaling factor applied to align an individual’s brain image to the Talairach atlas. This value is derived during the affine registration step, which scales the individual’s brain to match the atlas template. ASF is used to adjust for individual head size differences when calculating normalized volumes such as nWBV. eTIV is calculated using the inverse of the ASF, based on the assumption that the Talairach atlas has a known intracranial volume. Thus, eTIV = (Reference ICV) / ASF. This approach allows estimation of intracranial volume without direct segmentation. nWBV is derived by dividing the segmented whole brain tissue volume (gray and white matter) by the eTIV, providing a normalized measure of brain volume that accounts for inter-subject variability in head size. All registrations in this pipeline utilize affine transformation rather than nonlinear warping to preserve interpretability and consistency across individuals.

The dementia dataset, comprising clinical and demographic information for 150 unique individuals, reveals insightful patterns when analyzed by gender and age group. Stratification by gender shows that among the filtered 128 valid cases, males exhibit a higher proportion of dementia diagnoses—approximately 62% of males were diagnosed compared to just 36% of females, despite females forming a larger share of the total cohort. When data is grouped into 5-year age brackets from 60 to 94 years, a noticeable trend emerges: the number of dementia cases increases with age, peaking in the 75–79 age group and maintaining elevated levels into the 80s. Interestingly, both dementia and non-dementia cases are fairly balanced within the older age groups (80–84), indicating that while age is a critical factor, it is not solely determinative. The stacked histograms illustrate these gender and age-related disparities effectively, highlighting the need for nuanced, age- and gender-specific approaches to understanding and addressing dementia risk. **Figure 1** highlights that dementia prevalence increases with age and is notably higher among ages between 70∼84.

**Figure 1.**
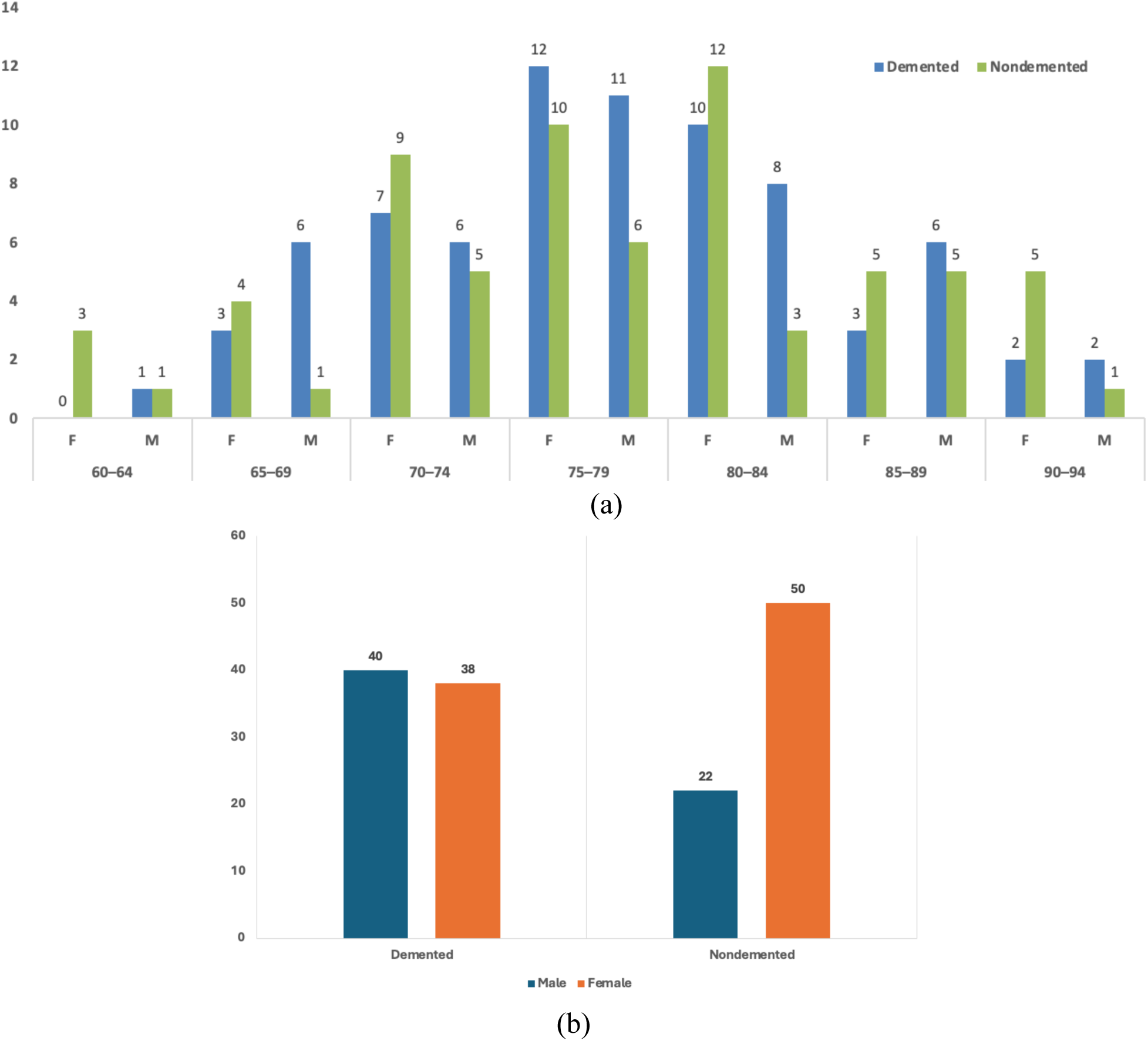
Distribution of dementia and non-dementia by age group and gender.

Based on the available dataset, this study aims to explore the potential for early symptom detection of dementia using machine learning techniques. By leveraging a combination of demographic, clinical, and neuroanatomical variables—such as age, MMSE scores, brain volume metrics (eTIV, nWBV), and education level—we aim to develop predictive models that accurately identify individuals at risk of dementia before severe cognitive decline. The integration of machine learning enables the uncovering of complex, non-linear relationships among variables that traditional statistical methods might overlook. This data-driven approach not only holds promise for improving diagnostic accuracy but also paves the way for proactive interventions, ultimately enhancing patient outcomes and reducing the long-term burden of dementia on individuals and healthcare systems.

### Data preprocessing

Before training selected models, it’s crucial to transform raw data into a clean, consistent, and suitable format. The process starts with exploring the dataset, understanding the structure, and identifying any missing values. The SES and MMSE variables each had missing entries, 19 and 2, respectively. MMSE is an essential variable in dementia diagnosis, as it measures an individual’s cognitive function across five areas: orientation, registration, attention and calculation, recall, and language [20]. The SES variable measures individuals’ socioeconomic status. Research shows that individuals with higher socioeconomic status received earlier diagnosis than others [21]. To address this, an advanced imputation method was deployed using R software and the missRanger package [22]. missRanger provides a fast, efficient way to impute missing values in a dataset using chained random forests. The algorithm predicts and fills in missing values by learning the relationships among all variables in the dataset.

**Figure 2** is a missing-data map that shows the completeness of each variable across all observations in the dataset. The gray areas represent present (non-missing) data, while the black segments highlight missing values. According to the figure, the overall data completeness is very high at 99.6%, with only 0.4% of values missing. Most variables—including Subject ID, MRI ID, Group, Visit, MR Delay, M/F, Hand, Age, EDUC, CDR, eTIV, nWBV, and ASF—are fully complete, with 0% missing values. However, a small proportion of missing data exists in the SES (5%) and MMSE (1%) variables. These minor gaps are unlikely to severely impact the analysis but should be addressed through imputation or case-wise exclusion, depending on the modeling approach. The figure provides a clear, concise quality check for the dataset, supporting its suitability for robust statistical and machine-learning applications.

**Figure 2.**
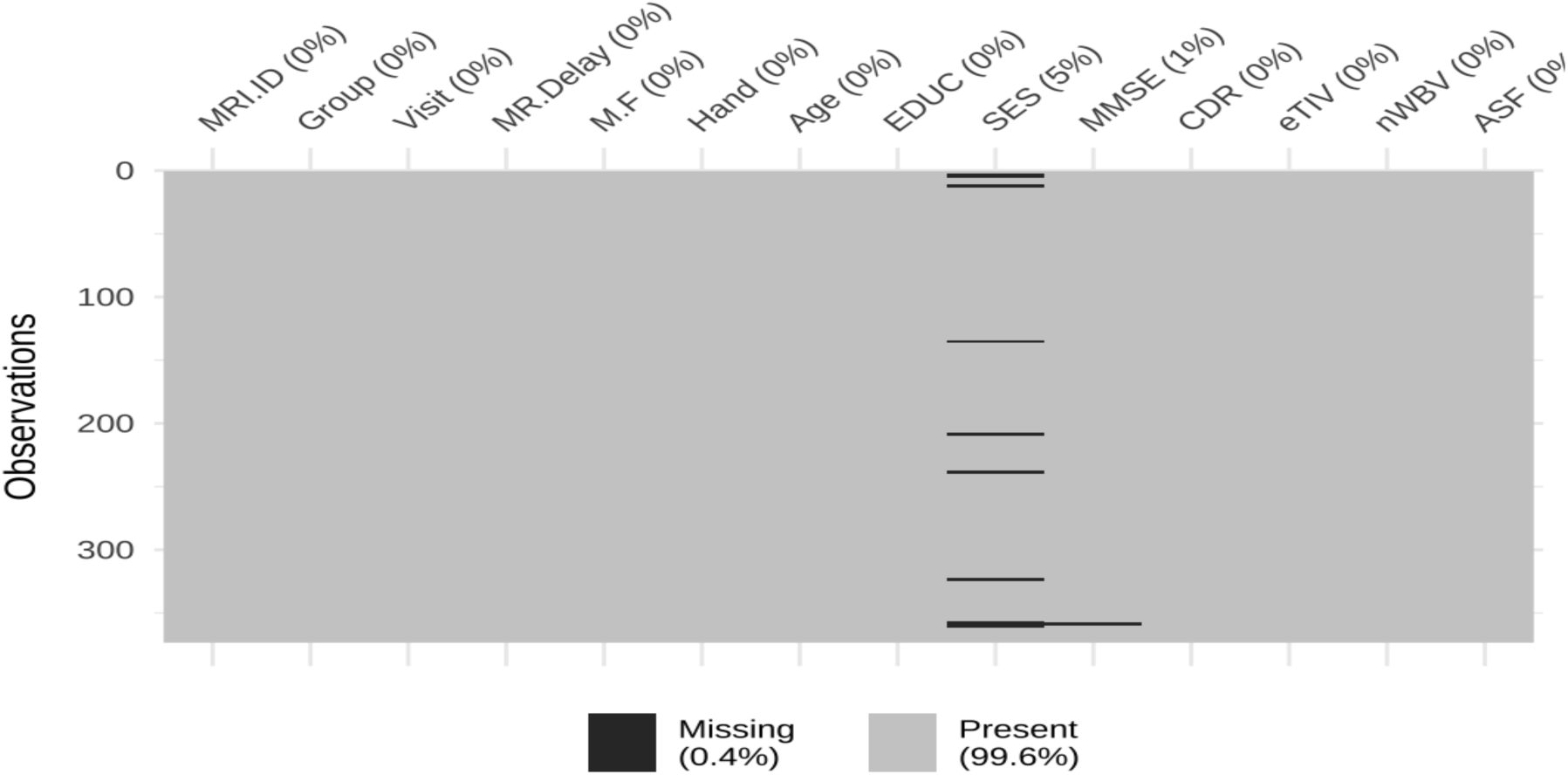
Heatmap of missing values in the original dataset.

The next step after addressing missing values is feature encoding, where categorical variables such as M/F, Hand, and Group are encoded as binary values. First, M/F is encoded so that “F” (females) is converted to 0 and “M” (males) is converted to 1. Next, all hand values are encoded: “R” (right-handed) is encoded as 1, and the dataset doesn’t include any left-handed individuals. Lastly, the Group variable containing three categories, “Nondemented”, “Demented”, and “Converted”, is encoded in binary as well. 14 individuals, about 9.91% of 373 scans, were “Converted” from “Non-demented”. The “Nondemented” individuals are encoded as 0, while both the “Demented” and “Converted” are encoded as 1. The “Converted” are encoded as 1 because over the course of the scans, they were diagnosed with dementia.

Here, individuals in the “Converted” group were classified as demented based on their final clinical diagnosis, consistent with established practice in longitudinal dementia studies. Specifically, subjects initially categorized as nondemented but later diagnosed with dementia were reassigned to the demented group using their last available visit, ensuring that each subject contributes a single, temporally consistent label. This approach avoids ambiguity associated with intermediate disease states and aligns with prior work using the OASIS dataset, where final diagnostic status is commonly used for classification analyses. This strategy is supported by the literature, which emphasizes the importance of using clinically confirmed endpoint diagnoses when transforming longitudinal data into cross-sectional frameworks. For example, DS Marcus et al. [32] and subsequent studies utilizing OASIS data adopt a similar approach to ensure consistent labeling and avoid misclassification of pre-conversion observations.

Moreover, as shown in the Figure 1(b), the cohort demonstrates a balanced class distribution across both groups. Among individuals classified as demented, 40 were male and 38 were female, whereas in the nondemented group, 22 were male and 50 were female. Overall, this corresponds to 78 demented cases (≈48.5%) and 72 nondemented cases (≈51.5%), indicating a relatively even distribution between outcome classes. This balanced representation reduces the likelihood of classification bias and supports stable model performance.

The final stage of preprocessing involves preparing the dataset for modeling. This consists of removing variables that lack predictive value, such as “Subject.ID”, “MRI.ID”, and “Hand”. The “Hand” variable, which contains only right-handed individuals, is removed from the dataset because it is irrelevant for predicting dementia status.

### Learning the data model and sampling

Before getting into the modelling part, we looked into the sample size of this study. Since this analysis is a cross-sectional analytical study designed to examine dementia status, with adequate sample size to ensure sufficient statistical power. Sample size estimation was performed using G*Power (version 3.1.9.4) for a χ² test of association between categorical variables. Assuming a 2-sided α of 0.05, an effect size (Cohen w) of 0.30, and 80% power, a minimum of 88 participants was required. The final analytic sample included 150 subjects, exceeding the prespecified requirement and supporting adequate statistical power for the planned analyses.

The dementia dataset was split into training and test sets to enable robust model development and evaluation. An 80/20 split ratio was employed using stratified sampling, with 80% of the data allocated to the training set and the remaining 20% to the testing set [23]. While stratified sampling is often preferred to maintain the original distribution of outcome classes across both subsets, it was not feasible in this case because at least one class had only a single observation, rendering stratification statistically invalid. Consequently, a randomized split without stratification was performed to ensure a fair and unbiased division of the data. Before splitting, the dataset was shuffled to minimize ordering bias. The training set was subsequently used to train machine learning models, while the unseen testing set provided an independent means to assess model generalization. This approach ensures that model performance metrics reflect dementia dataset scenarios in which predictions must be made on new, previously unencountered data.

### Machine Learning Model Development and Validation

To investigate the predictive outcome of the data, we analyzed 150 subjects and 373 MRI scans, each measuring multiple variables relevant to the condition under investigation. Out of 373 scans, some are repeated for the same subject; therefore, we consider the final visit for each patient as the final scan for that subject. To ensure a robust assessment of machine learning model performance, we adopted a 5-fold cross-validation procedure as shown in **Figure 3**. Also note that we used the cross-validation on the final data sets where there are 150 patients. This validation strategy is widely recognized for its effectiveness in minimizing overfitting and ensuring that the reported performance metrics are generalizable to new, unseen data. Missing values in clinical and neuroimaging variables are imputed using a chained random forest method, in which each variable with missing values is predicted from the remaining observed features. Categorical variables are encoded into binary indicators, and irrelevant identifiers such as subject ID, MRI ID, and handedness are removed. For feature selection, we use Random Forest importance scores based on the average reduction in Gini impurity across multiple trees, retaining the top 7 variables. These selected features are then used in a variety of classifiers, including Logistic Regression (LR), Decision Tree (CART), Random Forest, Gradient Boosting Machine (GBM), Xgboost, Multi-layer Perceptron (MLP), k-Nearest Neighbors (kNN), and Support Vector Machine (SVM). Each model is mathematically defined by its learning rule, ranging from logistic regression’s sigmoid transformation to SVM’s margin maximization. During 5-fold cross-validation, the entire dataset was randomly divided into five equal-sized, non-overlapping subsets. In each iteration, four folds were used to train the predictive model, while the remaining fold was used for validation. This process was repeated five times, with each fold serving as the validation set. This ensured that every data point was used exactly once for validation and four times for training. Following each iteration, several evaluation metrics were computed to quantify model performance on the validation data. The final reported metrics were then averaged across the five folds to account for variance due to different train-test splits. The primary metrics used to evaluate the models were Accuracy, Precision, Recall, F₁ Score, and the Matthews Correlation Coefficient (MCC). Each of these metrics provides unique insights into the classification performance and helps to form a comprehensive understanding of model behavior, especially when dealing with imbalanced datasets or medically significant features. Accuracy, calculated as 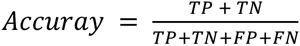, measures the overall proportion of correctly classified instances. While accuracy is intuitive and useful as a general indicator, it may be misleading when class distributions are imbalanced, which is often the case in medical imaging datasets. For this reason, we complemented accuracy with additional metrics. Precision, defined as 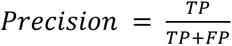 the proportion of true positives among all optimistic predictions made by the model, reflects the model’s ability to identify true positives correctly. High precision is critical in clinical applications, where false positives can lead to unnecessary interventions or anxiety. Recall (sensitivity), computed as the ratio of correctly identified actual positives to the total number of actual positives, indicates the model’s ability to identify all actual positives correctly. This metric is critical in medical diagnostics, where missing a positive case (i.e., a false negative) can have serious consequences. The F₁ score, the harmonic mean of precision and recall, balances these two metrics and is particularly useful when the trade-off between accuracy and recall must be optimized. It is sensitive to both false positives and false negatives, offering a more balanced view of model performance. Moreover, the Matthews Correlation Coefficient (MCC) is a robust metric for binary classification that accounts for true and false predictions across both classes. Unlike accuracy, MCC remains reliable even when class distribution is imbalanced, offering a more complete performance snapshot. MCC is calculated by 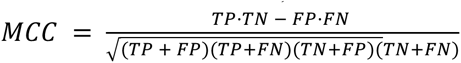. Note that the evaluation of these matrices is based on the confusion matrix derived from ML algorithms. Note that in classification tasks, a True Positive (TP) occurs when the model correctly identifies a data point as belonging to the positive class. At the same time, a True Negative (TN) indicates that the model correctly classifies a data point as belonging to the negative class. A False Positive (FP) arises when the model incorrectly labels a negative instance as positive, and a False Negative (FN) happens when the model fails to detect a positive instance, incorrectly classifying it as negative.

**Figure 3.**
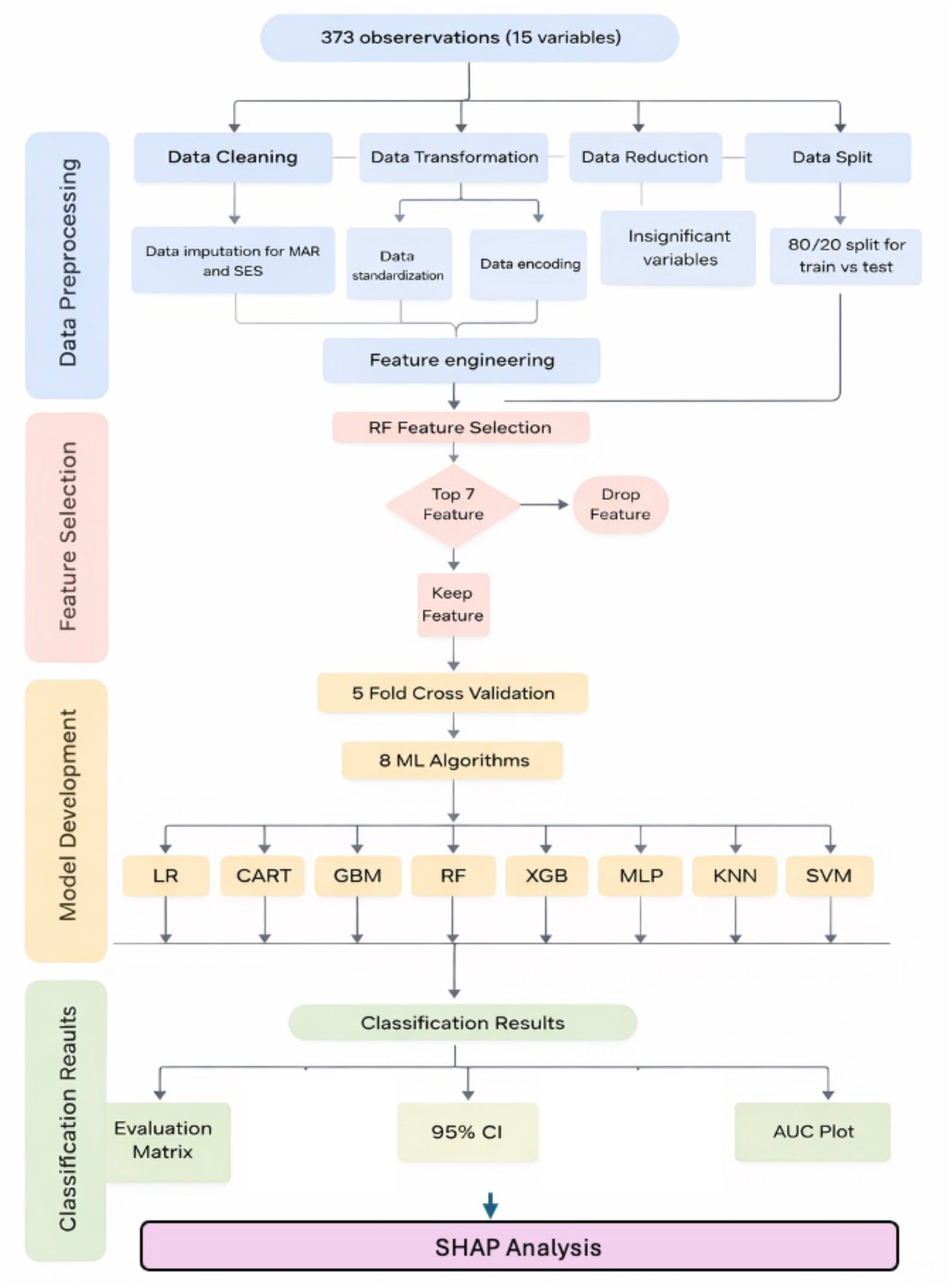
A streamlined ML pipeline showing data preprocessing, RF-based feature selection, model training with ML algorithms, and evaluation using metrics like MCC, AUC, and F1 score with 95% CI and AUC plots.

Across the entire dataset, 150 observations were used for the 7 top variables selected by the Random Forests feature selection algorithm used to develop the model. Comparison of the eight machine learning models yielded a detailed, well-informed landscape of their predictive abilities for distinguishing dementia. The performance of each model was extensively examined using a series of standard evaluation measures, providing a multifaceted assessment of its capabilities and shortcomings. The key performance indicators, for example, AUC, Accuracy, Sensitivity, Specificity, Precision, F₁-score, and MCC, are explained above and serve as the basis for the ensuing detailed analysis. The consistent high specificity across most models is particularly encouraging, as it indicates a low rate of false positives. In a clinical setting, this is crucial for avoiding unnecessary anxiety and further invasive testing for individuals who are not, in fact, on the path to dementia.

Moreover, it is important to note that, the validation pipeline was designed to ensure a robust and unbiased assessment of model performance while minimizing the risk of overfitting. Initially, the dataset was divided into an 80/20 train–test split, where 80% of the data was used for model development and the remaining 20% was held out as an independent test set with random seed state 42. This test set was not used at any stage of model training or tuning and served solely for final evaluation, providing an unbiased estimate of real-world performance. Within the training set, a 5-fold cross-validation procedure was applied to optimize model performance and assess stability. In this process, the training data was partitioned into five subsets, where four folds were used for training and one-fold for validation, iteratively across all folds. Importantly, all preprocessing steps, including feature selection, were performed within each training fold, ensuring that no information from the validation fold leaked into the training process. This two-stage validation approach combines the strengths of both cross-validation and independent testing. Cross-validation improves reliability by reducing variance in performance estimates, while the held-out test set provides a final, unbiased evaluation. Together, this framework ensures that the reported results are both stable and generalizable, addressing concerns related to overfitting and optimistic performance reporting.

Moreover, in this study, SHAP [33] is used to identify which features contribute most to dementia classification and to quantify both the magnitude and direction of each feature’s impact on the model output. This enhances transparency by showing, for example, how higher CDR or lower MMSE values increase the likelihood of a dementia prediction. Additionally, SHAP provides both global interpretability (overall feature importance) and local interpretability (individual-level explanations), which is critical in clinical settings where understanding the reasoning behind predictions is essential for trust, validation, and potential decision support.

### Computing platform, packages and study design

For this study, R was used exclusively for all computational tasks. To perform analysis, such as data preprocessing, the dlookr, naniar, missRanger, and corrplot packages were used. The dlookr package is used for data diagnosis and transformation to help assess data quality early in the analysis. The naniar package specializes in the exploration and visualization of missing data to help understand missing-data patterns before performing data imputation. The missRanger package is an accurate way to impute missing values in a dataset using the Random Forest algorithm. The corrplot package provides a way to visualize correlation matrices, which help understand the relationships between variables. For the machine learning tasks, the following packages were used: caret, randomForest, gbm, xgboost, nnet, kernlab, and e1071. These packages allow us to perform classification on the machine learning algorithms. Finally, to visualize and evaluate the results, ggplot2, pROC, and mltools were used. ggplot2 is the primary package for handling publication-quality graphics in R and was used to generate the ROC curve plots. Next, pROC was used to analyze the ROC curves and calculate the AUC and its confidence intervals. mltools is a collection of tools for performing machine learning tasks in R and was used to calculate the MCC score. In this study, random state = 42 set as a fixed seed and hyperparameters are presented in a table in the results section for reproduction.

## RESULTS

**Table 2** summarizes the clinical characteristics of individuals categorized by dementia status, offering insight into the statistical differences between those with and without the condition. For categorical variables (e.g., gender, SES, education), differences between groups were assessed using the Chi-squared test, while continuous variables (e.g., age, MMSE, CDR, brain volume measures) were analyzed using the Mann–Whitney U test due to potential non-normal distributions. Significant differences were observed in key clinical measures: individuals with dementia exhibited notably lower MMSE scores (median: 25 vs. 29) and higher CDR scores (median: 0.5 vs. 0.0), both with p-values < 0.001, reflecting the expected cognitive decline associated with dementia.

**Table 2.** Clinical characteristics by dementia status.

| Variable | No Dementia | Yes Dementia | P-value |
| --- | --- | --- | --- |
| Gender = F | 50 | 28 | 0.004 |
| Gender = M | 22 | 36 | 0.004 |
| SES = 1.0 | 15 | 11 | 0.273 |
| SES = 2.0 | 27 | 12 | 0.273 |
| SES = 3.0 | 16 | 15 | 0.273 |
| SES = 4.0 | 13 | 16 | 0.273 |
| SES = 5.0 | 1 | 2 | 0.273 |
| EDUC = 6 | 0 | 1 | 0.054 |
| EDUC = 8 | 1 | 3 | 0.054 |
| EDUC = 11 | 3 | 2 | 0.054 |
| EDUC = 12 | 13 | 26 | 0.054 |
| EDUC = 13 | 7 | 3 | 0.054 |
| EDUC = 14 | 6 | 5 | 0.054 |
| EDUC = 15 | 3 | 4 | 0.054 |
| EDUC = 16 | 16 | 11 | 0.054 |
| EDUC = 17 | 2 | 1 | 0.054 |
| EDUC = 18 | 19 | 5 | 0.054 |
| EDUC = 20 | 1 | 3 | 0.054 |
| EDUC = 23 | 1 | 0 | 0.054 |
| Age | 78.00 (73.00-84.25) | 77.50 (72.75-82.00) | 0.319 |
| MMSE | 29.00 (29.00-30.00) | 25.00 (21.00-28.00) | 0.000 |
| CDR | 0.00 | 0.50 (0.50-1.00) | 0.000 |
| eTIV | 1473.00 (1347.75-1599.50) | 1481.00 (1368.25-1563.75) | 0.846 |
| nWBV | 0.73 (0.71-0.76) | 0.70 (0.69-0.73) | 0.000 |
| ASF | 1.19 (1.10-1.30) | 1.19 (1.12-1.28) | 0.838 |

Furthermore, after applying the Benjamini–Hochberg false discovery rate (FDR) [35] correction for multiple comparisons, only the key clinical and neurocognitive variables such as MMSE, CDR, and nWBV remained statistically significant (adjusted p < 0.05), while other variables such as SES and EDUC were no longer significant. This indicates that cognitive performance and brain volume measures are the most robust discriminators between demented and non-demented groups. The findings reinforce the clinical relevance of these variables, as lower MMSE scores, higher CDR values, and reduced nWBV are strongly associated with dementia. Overall, the corrected results strengthen the validity of the conclusions by reducing false positives and highlighting the most reliable predictors in the dataset; however, further investigation using Random Forest–based feature importance is warranted to more comprehensively justify the selection of features for dementia prediction.

**Table 3** presents a comparative summary of key demographic and clinical characteristics between individuals classified as non-demented and those with dementia, with the "Converted" group included under the demented category. The data highlight distinct clinical differences between the two groups. The average MMSE score was substantially lower in the demented group (24.52 ± 4.74) compared to the non-demented group (29.17 ± 0.98), reflecting impaired cognitive function. Similarly, the Clinical Dementia Rating (CDR) was markedly higher among demented individuals. Although the mean age and estimated total intracranial volume (eTIV) were relatively comparable between the groups, normalized whole brain volume (nWBV) was lower in the demented group, consistent with brain atrophy. The gender distribution also shifted, with a higher proportion of females in the non-demented group (69.4%) than in the demented group (48.7%). These patterns underscore the clinical and structural brain differences associated with dementia and support the relevance of neurocognitive and volumetric markers in distinguishing dementia status.

**Table 3.** Demographic and clinical characteristics by dementia status after adding the converted to dementia group.

| Variable | No Dementia | Demented |
| --- | --- | --- |
| Number | 72 | 78 |
| Age (Mean ± SD) | 78 ± 8 | 78 ± 7 |
| MMSE (Mean ± SD) | 29.17 ± 0.98 | 24.52 ± 4.74 |
| CDR (Mean ± SD) | 0.00 ± 0.00 | 0.69 ± 0.35 |
| eTIV (Mean ± SD) | 1488.43 ± 191.59 | 1481.74 ± 167.86 |
| nWBV (Mean ± SD) | 0.734 ± 0.039 | 0.712 ± 0.035 |
| ASF (Mean ± SD) | 1.198 ± 0.150 | 1.199 ± 0.133 |

The correlation matrix in **Figure 4** provides insight into the pairwise linear relationships among various clinical and demographic variables, including EDUC, MMSE, nWBV, SES, ASF, M.F, CDR, Visit, and Age. Several relationships are noteworthy. The strongest negative correlation is between EDUC and SES (r = -0.72), suggesting that higher education levels are associated with lower SES scores—this might reflect how SES is coded (e.g., lower values representing higher status). Additionally, MMSE and CDR show a strong negative correlation (r = -0.69), indicating that as cognitive function declines (lower MMSE), the severity of dementia increases (higher CDR). nWBV correlates negatively with CDR (r = -0.34) and Age (r = -0.52), as expected, since brain volume typically decreases with age and dementia severity. The negative correlation between ASF and M.F (r = -0.56) may point to some technical or anatomical association, such as brain scaling inversely related to specific functional measurements.

**Figure 4.**
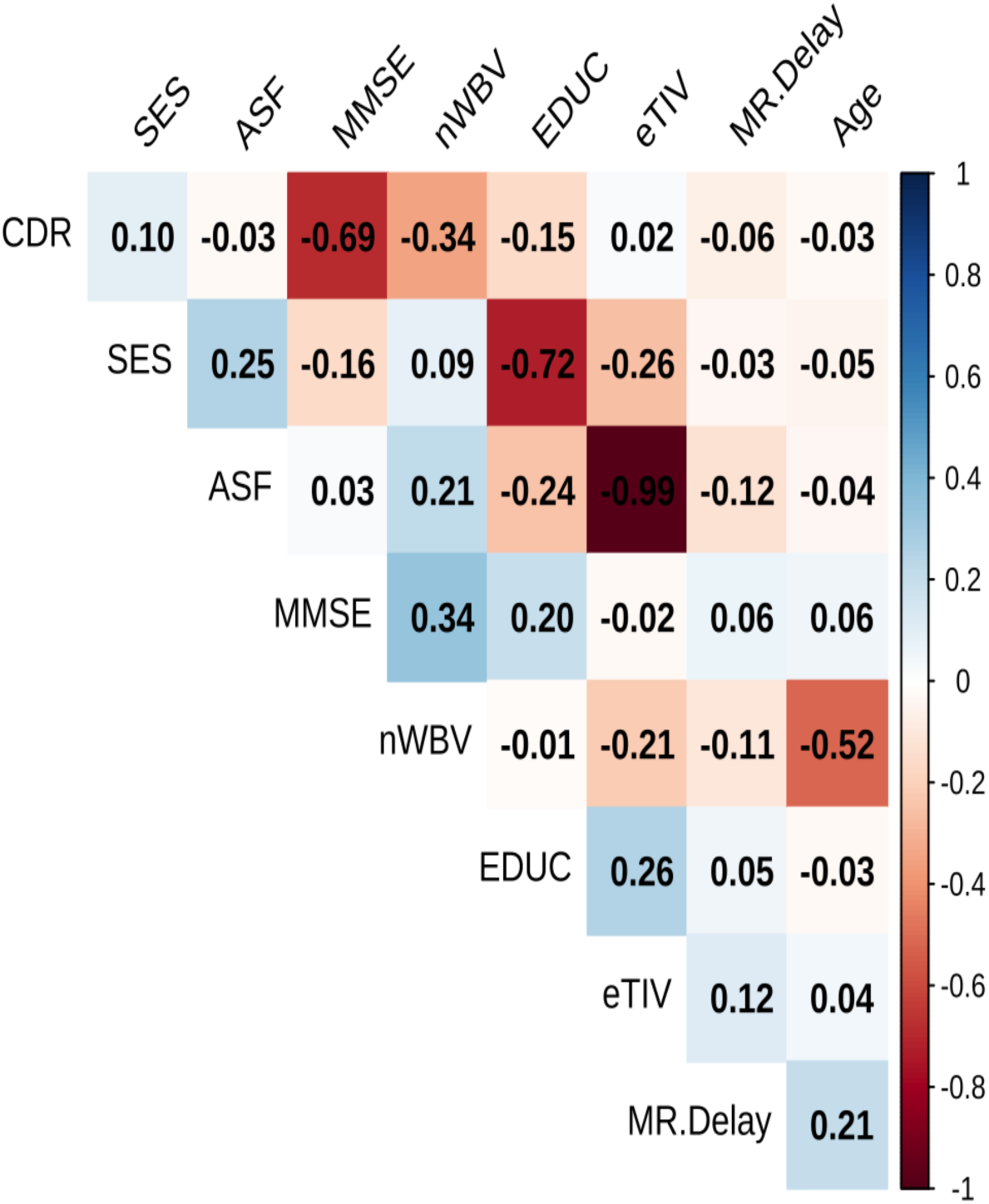
Heatmap of variable interrelationships in clinical, demographic, and imaging variables.

On the other hand, variables such as Visit, Age, and M.F show weak or negligible correlations with most other variables, suggesting low multicollinearity and relative independence in the dataset. For instance, Age is only moderately negatively correlated with nWBV and weakly with most other variables.

In terms of collinearity, no pair (except EDUC-SES and MMSE-CDR) exhibits very high correlation (|r| > 0.8), suggesting that multicollinearity is not a significant concern for regression modeling with this data. However, the strong EDUC–SES and MMSE–CDR relationships should be carefully managed to avoid redundancy and inflated variance in model coefficients.

Moreover, in **Figure 4**, a collinearity is between ASF and eTIV (r = -0.99), indicating that these two variables are nearly perfectly inversely correlated. Including both in a regression model would introduce redundancy and inflate standard errors. Therefore, one of these should be removed or carefully transformed before modeling. Similarly, Visit and MR Delay are highly positively correlated (r = 0.92), suggesting they represent similar temporal dimensions and possibly capture the same underlying timing variable. Retaining both would likely contribute to multicollinearity, so one should be excluded based on clinical relevance. Other correlations, while moderate, remain below the 0.75 threshold and do not warrant removal. After removing highly correlated par, **Figure 5** displays the correlation matrix among clinical, demographic, and imaging variables after removing highly correlated pairs (|r| > 0.75), revealing predominantly weak-to-moderate associations and limited multicollinearity, for example, MMSE and CDR are negatively correlated (r = -0.69), consistent with clinical expectations: lower cognitive scores are associated with higher dementia severity. nWBV and Age also show a moderate negative correlation (r = -0.52), indicating a decrease in brain volume with age. Importantly, EDUC and SES have a strong negative correlation (r = -0.72), approaching the collinearity threshold. This near redundancy implies overlapping socioeconomic information, and careful judgment is needed if both are to be included in predictive models. After removing the highly correlated variable pairs (ASF–eTIV, Visit–MR Delay), the matrix shows a relatively low level of multicollinearity among the remaining features. This reduced correlation set can enhance the interpretability and numerical stability of subsequent regression or machine learning models, ensuring that each variable contributes uniquely to the prediction task. In the next part we introduced the RF-based feature selection strategy to support the results from above correlation-based feature analysis.

**Figure 5.**
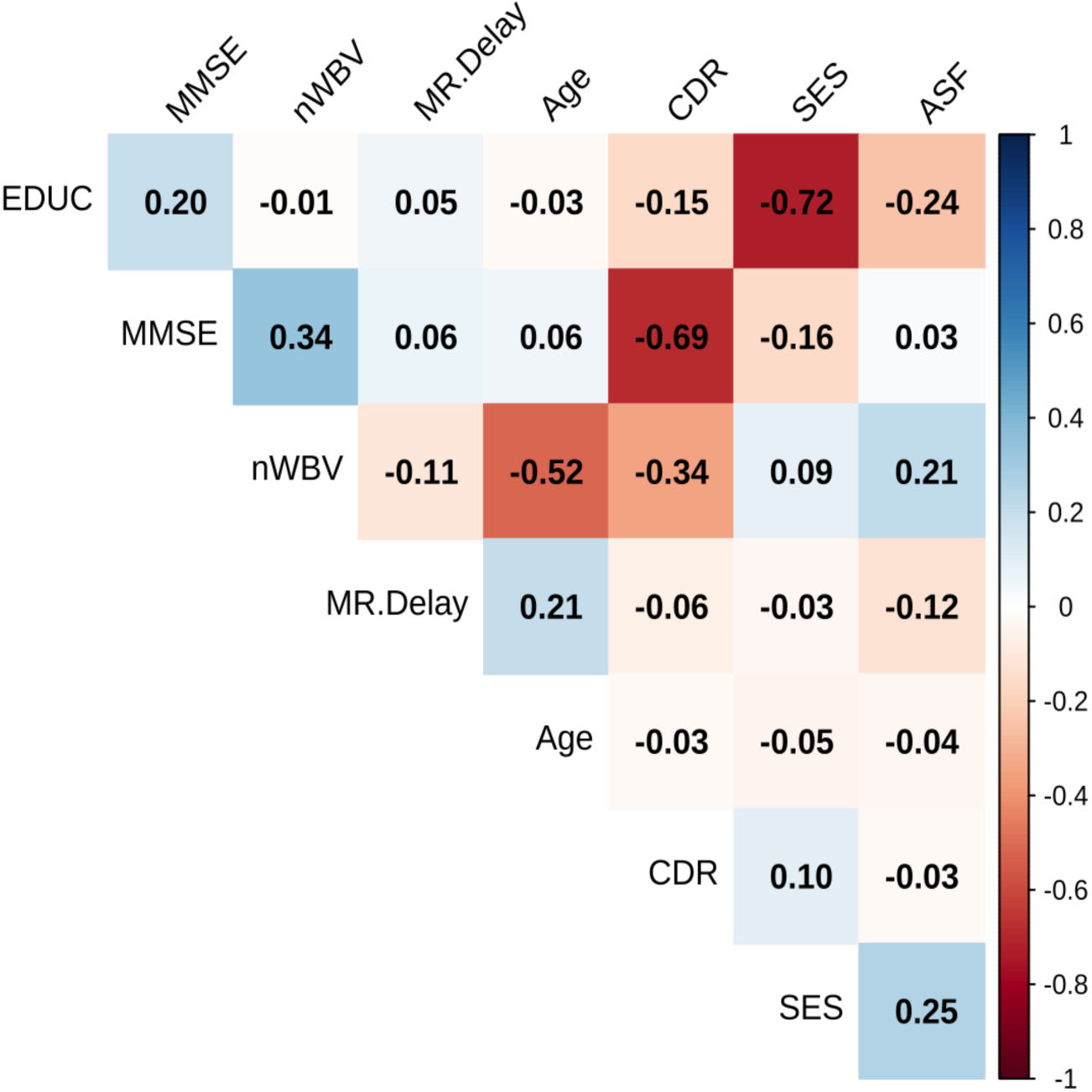
Correlation matrix of clinical, demographic, and imaging variables after removing highly correlated pairs (|r| > 0.75).

The Random Forest feature importance plot above provides valuable insights into which variables most strongly influence model predictions. A key strength of this approach lies in the implementation of RF-based feature selection within each cross-validation fold. By selecting features independently within each training fold, the model avoids information leakage and ensures that feature importance is derived solely from the training data. This nested-like validation strategy enhances the reliability of the reported performance metrics and provides a more realistic estimate of how the model would perform in real-world settings. The RF feature importance in **Figure 6** is derived using two complementary metrics: Mean Decrease in Accuracy (MDA) and Mean Decrease in Gini (MDG). The Mean Decrease in Accuracy quantifies the reduction in model performance when the values of a given feature are randomly permuted, thereby reflecting its contribution to predictive accuracy. In contrast, the Mean Decrease in Gini measures the total reduction in node impurity attributable to each feature across all trees in the forest, indicating its role in improving class separation during model training. As shown in **Figure 6**, CDR consistently ranks as the most important predictor under both metrics, followed by MMSE and structural brain measures such as nWBV and ASF, confirming their strong contribution to dementia classification. While both metrics provide similar rankings, MDA is more directly linked to model performance, whereas MDG reflects the internal splitting behavior of the model. In this study, feature selection was based on the ranking derived from RF importance (primarily guided by MDA) to ensure that selected features contribute meaningfully to predictive performance. This dual-metric approach improves interpretability and supports the robustness of the selected feature set.

**Figure 6.**
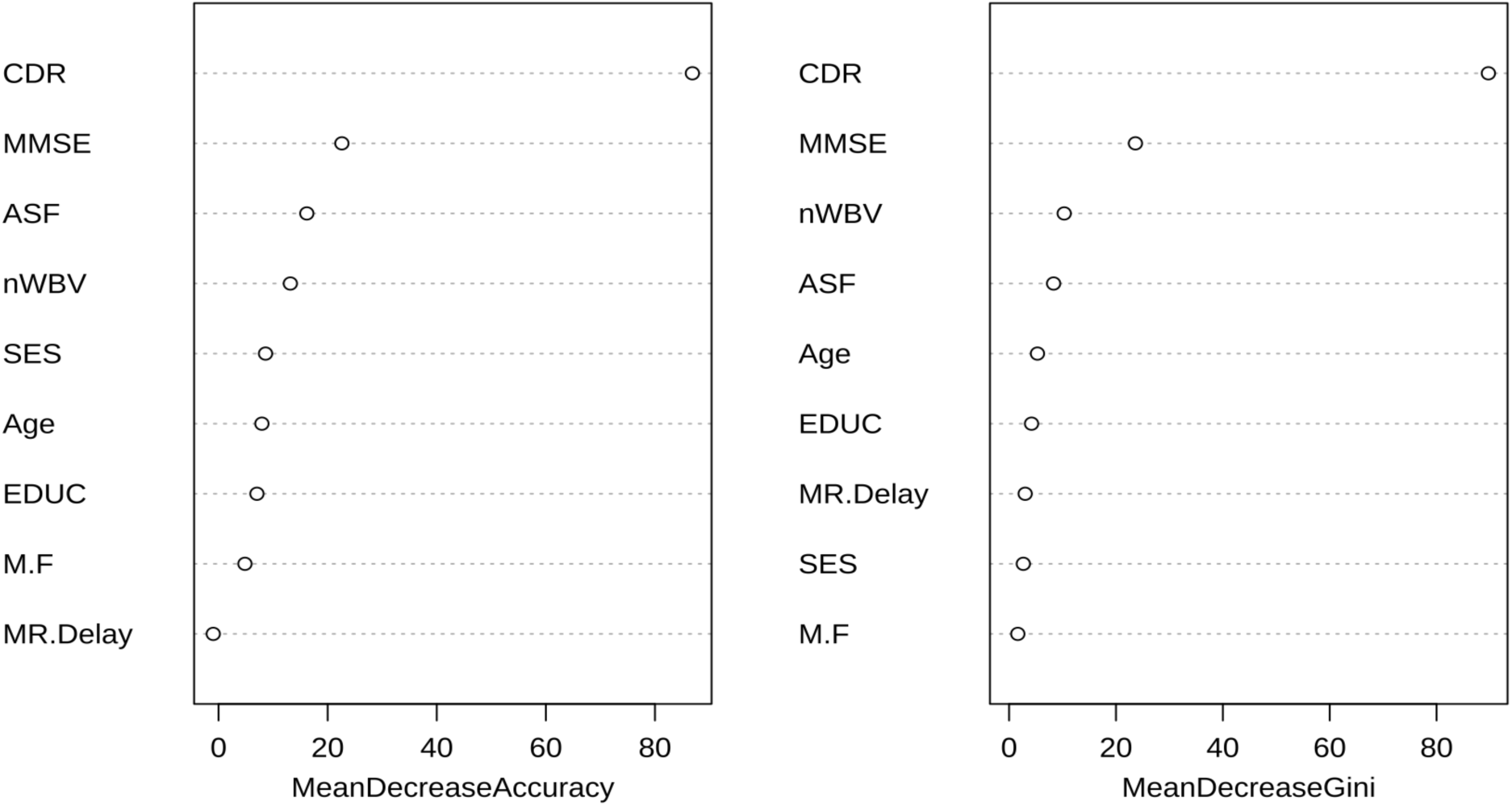
Ranking predictive features for dementia diagnosis based on RF feature selection.

Moreover, in this context, the model was applied to a clinical and neuroimaging dataset to assess the relative predictive value of features such as cognitive scores (e.g., MMSE), structural brain metrics (e.g., ASF, nWBV), demographics (e.g., Age, EDUC), and clinical indicators (e.g., CDR). Random Forest is an ensemble learning method that builds multiple decision trees and averages their outputs to improve prediction accuracy and control overfitting. One key advantage of this approach is its built-in mechanism to evaluate feature importance. This is typically done by measuring how much each variable reduces impurity (e.g., Gini impurity or entropy) at the trees’ internal nodes across the ensemble. Features that consistently result in significant impurity reductions across many trees are considered more important. In Figure 6, CDR emerges as the most influential feature by a large margin, indicating it plays a dominant role in the model’s decision-making process. This is expected, as CDR captures overall dementia severity and likely correlates with both clinical and cognitive outcomes. MMSE follows next, aligning with its role as a standard measure of mental performance.

More importantly, CDR evaluates the severity of dementia based on functional performance in domains such as memory, orientation, and judgment, while MMSE provides a quantitative score of global cognitive function, including memory, attention, and language abilities. Moreover, in this study, the inclusion of CDR and MMSE was intended to provide clinically meaningful baseline cognitive information and to improve model interpretability by aligning predictions with established diagnostic markers. Importantly, the model does not rely solely on these variables but integrates them with neuroimaging (e.g., nWBV) and demographic features, allowing it to capture multidimensional patterns beyond clinical scoring alone. Therefore, in this context based on feature selection model performance, both features are used since MMSE serves as a rapid cognitive screening tool that can approximate dementia severity and act as a practical surrogate in clinical settings [31].

Other essential features include ASF, SES, and EDUC, reflecting how brain size normalization, social determinants, and educational background contribute meaningfully to model predictions. Notably, visit has near-zero importance, suggesting it provides little discriminative information in this model—likely due to high correlation with other time-related variables, such as MR delay, or to the redundancy of visit counts across patients. This ranking helps prioritize variables for model simplification, clinical interpretation, or further validation. It also helps mitigate overfitting by allowing the exclusion of low-importance features, enhancing the model’s generalizability and interpretability. Overall, Random Forest feature importance provides a transparent and data-driven method for identifying key predictors in complex clinical datasets. **Table 4** summarizes the key hyperparameters tuned for each machine learning model considered in our analysis. While logistic regression requires no tuning, models such as CART, k-nearest neighbors, and support vector machines rely on parameters controlling complexity, neighborhood size, and kernel behavior. Ensemble methods like random forests, gradient boosting, and XGBoost involve choices about tree depth, the number of trees, and learning rates, reflecting the trade-off between bias and variance. Finally, for neural networks, the hidden layer size and weight decay govern model capacity and regularization, respectively. Together, these hyperparameter grids ensure a systematic exploration of model performance across diverse algorithmic families.

**Table 4.** Machine learning models and their corresponding hyperparameters with candidate values considered for tuning during model selection.

| Model (R function) | Hyperparameter(s) | Values Tested |
| --- | --- | --- |
| Logistic Regression (glm) | None | N/A |
| CART (rpart) | cp (Complexity Parameter) | seq (0.001, 0.05, by = 0.005) |
| K-Nearest Neighbors (knn) | k (Number of Neighbors) | seq (1, 19, by = 2) |
| Support Vector Machine (svmRadial) | C (Cost) | 0.1, 1, 10 |
|  | sigma (Kernel Scale) | 0.01, 0.1, 1 |
| Random Forest (rf) | mtry (# variables per split) | 2, 3, 4, 5 |
| Gradient Boosting (gbm) | n.trees (# Trees) | 100, 150 |
|  | interaction.depth | 1, 2, 3 |
|  | shrinkage (Learning Rate) | 0.01, 0.1 |
| XGBoost (xgbTree) | nrounds (# Rounds) | 100, 150 |
|  | max_depth | 3, 5 |
|  | eta (Learning Rate) | 0.1, 0.3 |
| Multilayer Perceptron (nnet) | size (# Hidden Units) | 3, 5, 7 |
|  | decay (Weight Decay) | 0.01, 0.1, 0.5 |

To get better performance, various machine learning models were optimized using key hyperparameters. For RF, the primary hyperparameters included the number of trees (500), maximum tree depth (4), and the number of features considered for splits (7). SVM was tuned using the regularization parameter (2), kernel type (RBF), and kernel coefficient (0.1). For GBM and XGB, important hyperparameters included the number of boosting rounds (1000), the learning rate (0.01), the maximum tree depth (4), and the subsampling ratio (0.5). kNN was adjusted based on the number of neighbors (7), distance metric (metric), and weight function (weights). The MLP used hidden_layer_sizes to define the network architecture, along with activation, solver, and learning_rate_init to control the learning process. For LR, the key hyperparameters were the inverse regularization strength (C) and the regularization penalty type (penalty). Lastly, CART was configured using max_depth, min_samples_split, and the splitting criterion (criterion), such as Gini or entropy.

These hyperparameters were selected to enhance predictive accuracy, generalizability, and model efficiency across all classifiers.

**Table 5** shows how different machine learning models perform in predicting dementia. Each model is evaluated to highlight its strengths and weaknesses. The models assessed include RF, SVM, GBM, XGboost, kNN, MLP, LR, and CART. Also, **Table 5** presents the performance metrics of multiple machine learning models on the training set for distinguishing between Demented and No Dementia cases. The metrics include Accuracy, Kappa, Sensitivity, Precision, and F1 score, each accompanied by confidence intervals. These results highlight the strengths and weaknesses of the different classifiers, providing insight into their learning behavior and potential clinical applications. Starting with Accuracy, ensemble methods such as RF and GBM achieved the highest mean accuracies (0.9960 and 0.9967, respectively), suggesting that these models capture complex patterns in the data exceptionally well. CART and Logistic Regression also performed strongly, with accuracies around 0.956–0.960, while KNN and MLP were slightly lower but still above 0.94. Notably, both XGBoost and SVM achieved or exceeded 0.986 in training accuracy, indicating consistent high training accuracy. The uniformly high performance across models may also reflect the possibility of overfitting, particularly for ensemble methods. The Kappa statistic, which adjusts for chance agreement, provides a more reliable measure of discriminative ability. RF again excelled, achieving a near-perfect Kappa (0.9950), demonstrating highly reliable classification beyond chance. GBM followed closely (0.9933), reinforcing its robustness. XGBoost and SVM both recorded strong Kappa values (0.9732), while MLP achieved 0.9464. By contrast, Logistic Regression and CART performed adequately (around 0.91), while KNN was the weakest at 0.8995, suggesting that simpler models may have limited agreement compared to ensemble approaches.

**Table 5.** Performance metrics of machine learning models on the training set for classifying demented versus no dementia subjects. Reported values include mean accuracy, Kappa, sensitivity, precision, and F1 score, with 95% confidence intervals in parentheses.

| Model | Accuracy | Kappa | Sensitivity | Precision | F1 |
| --- | --- | --- | --- | --- | --- |
| LogReg | 0.9565<br>(0.9334, 0.9796) | 0.9130<br>(0.8811, 0.9449) | 0.9320<br>(0.9035, 0.9605) | 0.9786<br>(0.9622, 0.9950) | 0.8750 (0.8376, 0.9124) |
| CART | 0.9599<br>(0.9377, 0.9821) | 0.9196<br>(0.8888, 0.9504) | 0.9320<br>(0.9035, 0.9605) | 0.9856<br>(0.9721, 0.9991) | 0.8750 (0.8376, 0.9124) |
| RF | 0.9960<br>(0.9889, 1.0000) | 0.9950<br>(0.9870, 1.0000) | 0.9960<br>(0.9889, 1.0000) | 0.9320<br>(0.9035, 0.9605) | 0.9320 (0.9035, 0.9605) |
| GBM | 0.9967<br>(0.9902, 1.0000) | 0.9933<br>(0.9841, 1.0000) | 0.9933<br>(0.9841, 1.0000) | 0.9320<br>(0.9035, 0.9605) | 0.9966 (0.9900, 1.0000) |
| XGBoost | 0.9866<br>(0.9736, 0.9996) | 0.9732<br>(0.9549, 0.9915) | 0.9796<br>(0.9636, 0.9956) | 0.9320<br>(0.9035, 0.9605) | 0.9863 (0.9731, 0.9995) |
| MLP | 0.9732<br>(0.9549, 0.9915) | 0.9464<br>(0.9209, 0.9719) | 0.9456<br>(0.9199, 0.9713) | 0.9320<br>(0.9035, 0.9605) | 0.9720 (0.9533, 0.9907) |
| KNN | 0.9498<br>(0.9251, 0.9745) | 0.8995<br>(0.8655, 0.9335) | 0.9048<br>(0.8716, 0.9380) | 0.9925<br>(0.9827, 1.0000) | 0.9466 (0.9212, 0.9720) |
| SVM | 0.9866<br>(0.9736, 0.9996) | 0.9732<br>(0.9549, 0.9915) | 0.9796<br>(0.9636, 0.9956) | 0.9931<br>(0.9837, 1.0000) | 0.9863 (0.9731, 0.9995) |

Sensitivity (recall), crucial in clinical contexts for identifying demented individuals, was highest for RF (0.9960) and GBM (0.9933), indicating that these models rarely miss dementia cases. CART and Logistic Regression reported lower but still respectable sensitivities (0.9320), while XGBoost (0.9796) and SVM (0.9796) also performed strongly. MLP had the highest sensitivity (0.9456), and KNN had the lowest (0.9048). High sensitivity is critical in dementia detection, as false negatives may delay diagnosis and treatment. Thus, RF and GBM stand out as the most clinically effective in minimizing missed cases.

On the other hand, precision reflects the reliability of optimistic predictions (i.e., the proportion of correctly identified Demented cases among all predicted positives). CART achieved the highest precision (0.9856), followed closely by Logistic Regression (0.9786), demonstrating that these models are particularly conservative in labeling subjects as demented, thereby minimizing false positives. SVM also stood out with a precision of 0.9931, nearly perfect in avoiding misclassifications. RF and GBM, despite their high sensitivity, showed lower precision (0.9320), suggesting a trade-off: these models capture more true positives but also introduce more false positives. KNN demonstrated the best precision overall (0.9925), though at the expense of lower sensitivity, highlighting its bias towards avoiding false alarms. The F1 score, a harmonic mean of precision and recall, provides a balanced measure. GBM excelled with an F1 score of 0.9966, reflecting both high sensitivity and substantial precision, making it the best all-around performer. RF followed with 0.9320, though its lower precision limited its F1 score. XGBoost and SVM both recorded impressive F1 values (0.9863), while MLP also performed well (0.9720). Logistic Regression and CART had relatively lower F1 scores (0.8750), mainly due to an imbalance between precision and sensitivity. Overall, the training set results suggest that GBM provides the most balanced and reliable classification of Demented vs. No Dementia, followed closely by RF, XGBoost, and SVM. Simpler models, such as Logistic Regression and CART, demonstrate substantial precision but weaker balance, while KNN struggles with consistency across metrics. While these results underscore the strength of ensemble methods, the exceptionally high performance on the training set warrants caution, as it may reflect overfitting and necessitate validation on test data to confirm generalizability.

Moreover, the cross-validation results indicate exceptionally high and consistent performance across nearly all machine learning models. Form **Table 6**, we can observe that, ML models such as RF, SVM, GBM, XGB, MLP, LR, and CART achieved almost identical performance, with accuracy (∼0.991), Kappa (∼0.983), sensitivity (∼0.982), and F1-score (∼0.990). The precision of 1.000 across models suggests no false positives during cross-validation, while AUC values approaching 1.000 demonstrate excellent discriminative ability. However, the near-perfect performance across multiple models raises concerns about potential overfitting, especially given the relatively small dataset. The kNN model, with slightly lower performance (accuracy 0.974, Kappa 0.948), provides a useful contrast, suggesting that not all models generalize equally well. Overall, while these results are promising, they emphasize the importance of independent test-set validation to confirm real-world applicability, which is discussed below.

**Table 6.** Cross-validated performance metrics of machine learning models on the for classifying demented versus no dementia subjects.

| Model | Accuracy | Kappa | Sensitivity | Precision | F1 Score | AUC |
| --- | --- | --- | --- | --- | --- | --- |
| RF | 0.991 | 0.983 | 0.982 | 1.000 | 0.990 | 1.000 |
| SVM | 0.991 | 0.983 | 0.982 | 1.000 | 0.990 | 0.998 |
| GBM | 0.991 | 0.983 | 0.982 | 1.000 | 0.990 | 0.991 |
| XGB | 0.991 | 0.983 | 0.982 | 1.000 | 0.990 | 0.991 |
| KNN | 0.974 | 0.948 | 0.947 | 1.000 | 0.971 | 0.999 |
| MLP | 0.991 | 0.983 | 0.982 | 1.000 | 0.990 | 1.000 |
| LogReg | 0.991 | 0.983 | 0.982 | 1.000 | 0.990 | 1.000 |
| CART | 0.991 | 0.983 | 0.982 | 1.000 | 0.990 | 0.991 |

The results from the cross-validation and independent test set demonstrate that the proposed modeling framework achieves robust and generalizable performance while effectively mitigating overfitting. The 5-fold cross-validation results, with mean accuracy and AUC values in the range of approximately 0.99–1.0, indicate that the model performs consistently across different subsets of the training data. This consistency suggests that the learned patterns are not driven by random fluctuations or specific partitions of the dataset.

Importantly, the independent test-set performance remains closely aligned with the cross-validation results, with only a slight decrease in accuracy and AUC. This minimal performance gap is a strong indicator that the model generalizes well to unseen data and is not overfitted to the training set. In many machine learning studies, a large discrepancy between training and testing performance signals overfitting; however, such a pattern is not observed here.

**Figure 7** presents a comprehensive comparison of several machine learning models for the classification task of distinguishing between Demented and No Dementia subjects from test sets, using key performance metrics—Accuracy, Kappa, Sensitivity, Precision, and F1 score—along with their respective error bounds. This multidimensional evaluation allows us to critically assess not only the central tendencies of each metric but also the variability and reliability of the estimates, which is crucial when models are applied to sensitive clinical decision-making problems such as dementia diagnosis. Starting with accuracy, most machine learning models demonstrated competitive performance. The mean accuracies generally ranged from approximately 0.86 to 0.91, indicating a strong overall ability to predict dementia across algorithms. Among these models, RF and GBM stood out slightly. They not only achieved higher accuracy scores but also maintained stability, as reflected in their relatively narrow error margins. This consistency suggests that RF and GBM offer more reliable predictions in practical applications. This indicates that ensemble-based approaches are particularly effective at capturing the complex, potentially nonlinear patterns in the clinical data that differentiate demented from non-demented individuals. By contrast, SVM and KNN yielded lower accuracies with broader error bounds, reflecting reduced robustness in this application.

**Figure 7.**
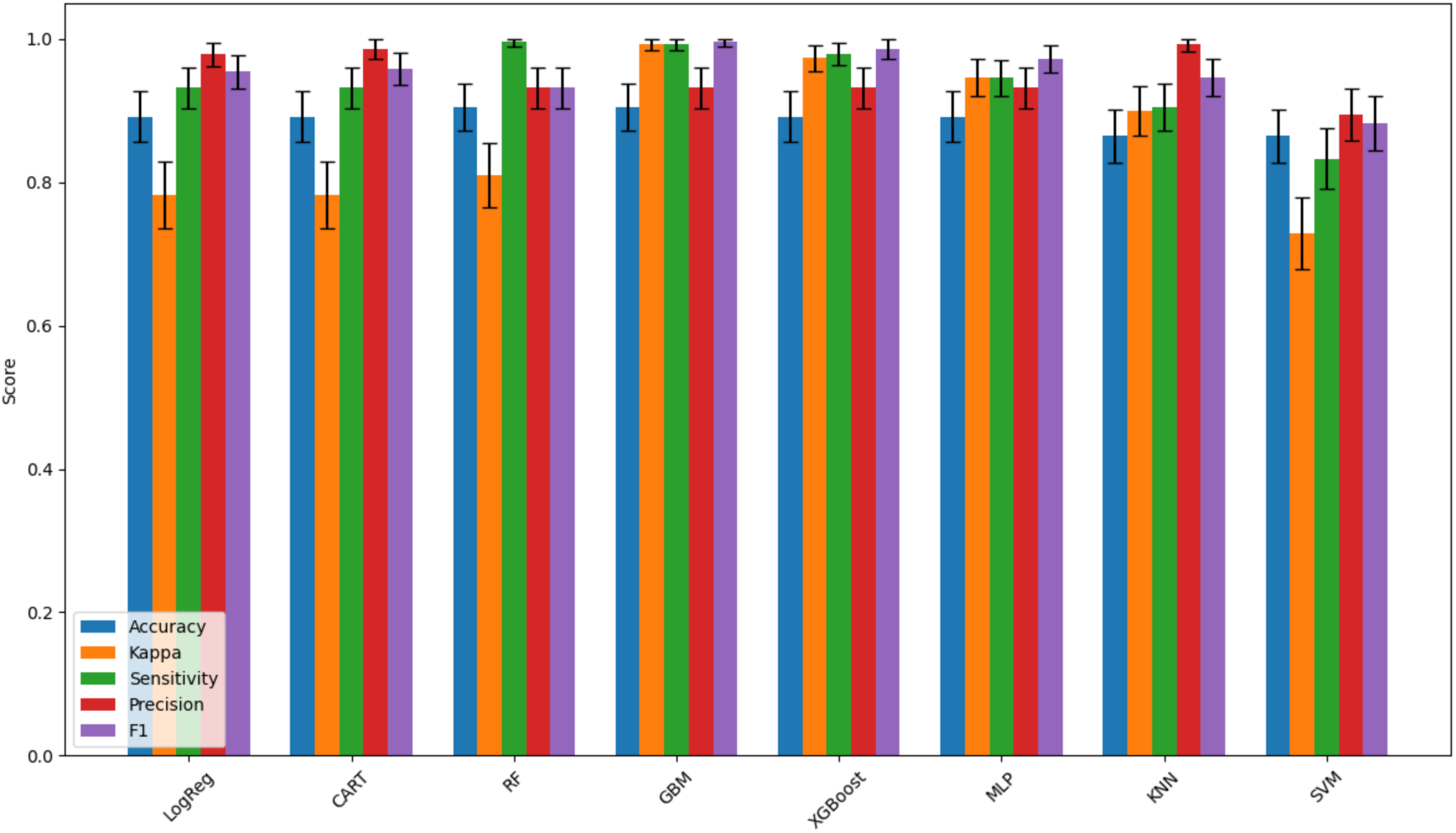
Comparison of machine learning models for distinguishing between Demented and No Dementia subjects using test sets. Bars represent mean performance across five metrics (Accuracy, Kappa, Sensitivity, Precision, and F1 score), with error bars indicating the corresponding confidence intervals. Ensemble methods, particularly GBM and RF, achieved the most balanced and reliable performance across metrics, highlighting their suitability for clinical dementia classification tasks.

The Kappa statistic, which accounts for chance agreement, provides additional insight. GBM achieved near-perfect Kappa values (close to 1.0), indicating excellent reliability in distinguishing between the two diagnostic groups beyond chance. XGBoost and MLP also demonstrated strong Kappa values, whereas LogReg and CART scored moderately, indicating more limited agreement. Interestingly, SVM produced the lowest Kappa, reinforcing its weaker performance and suggesting that its predictions may be less clinically reliable in this dataset. Sensitivity (recall), particularly critical in dementia screening because it reflects the ability to identify demented individuals correctly, was highest for RF and GBM, both approaching near-perfect detection rates. This outcome emphasizes the strength of ensemble methods in minimizing false negatives—an essential requirement in medical diagnostics, where failing to detect dementia could delay treatment and intervention. XGBoost also performed strongly on sensitivity, though slightly lower than GBM, while LogReg and CART were acceptable but less optimal. SVM lagged substantially, raising concerns about its suitability for screening tasks in this clinical setting. Precision, or positive predictive value, complements sensitivity by reflecting the proportion of correctly classified dementia cases among those predicted as demented. Both CART and KNN achieved high precision, approaching 1.0, indicating that their optimistic predictions are highly reliable. This is valuable for avoiding false alarms that may cause unnecessary anxiety for patients labeled as demented when they are not. However, these models did not achieve the same balance in sensitivity, highlighting the classical trade-off between recall and precision. The F1 score, which balances precision and sensitivity, provides a holistic assessment. GBM again led with the highest F1, indicating that it achieves both strong sensitivity and precision, an ideal characteristic in clinical screening models. XGBoost and MLP followed closely, while RF performed reasonably but showed slightly lower precision despite high sensitivity. SVM and KNN, by contrast, exhibited weaker balance, making them less suitable for general use in dementia classification. Overall, the figure underscores the superiority of ensemble methods, particularly RF, for classifying Demented vs. No Dementia. Overall, this suggests that tree-based ensemble learning method RF offers a robust framework for dementia detection and could serve as a robust decision-support tool in medical practice. These results provide a strong foundation for future research, including prospective validation of these models in clinical trials and their potential integration into decision-support systems for clinicians.

Moreover, in the supplementary file **Table 7** summarizes model performance across three feature scenarios. Models using both CDR and MMSE demonstrated near-perfect discrimination, with cross-validation accuracy of 0.99 and AUC of 1.00, which was maintained in the independent test set. When CDR was excluded, performance declined substantially (e.g., RF cross-validation accuracy 0.74; AUC 0.82; SVM accuracy 0.81; AUC 0.86), indicating reduced discriminatory ability. In contrast, exclusion of MMSE resulted in minimal change, with performance remaining high (RF cross-validation accuracy ≥0.98 and AUC ≥0.99). These findings were consistent across cross-validation and test sets, supporting the dominant contribution of CDR to model performance relative to MMSE.

The ROC curves in **Figure 8** panels (a), (b), and (c) illustrate how varying model configurations—specifically, different feature subsets derived through Random Forest–based feature selection (top 3, top 5, and top 7 features)—as well as classifier settings and preprocessing strategies, affect the sensitivity–specificity trade-off in dementia classification. Each ROC curve plots the True Positive Rate (Sensitivity) against the False Positive Rate (1 − Specificity) across multiple decision thresholds, providing both a visual and quantitative comparison of classifier performance. The Area Under the Curve (AUC) serves as a summary measure of each model’s ability to discriminate between demented and nondemented individuals, with higher AUC values indicating stronger discriminatory capacity. Importantly, feature subsets were determined within the training data using Random Forest feature importance and evaluated under the same validation framework, ensuring that comparisons across panels reflect differences attributable to feature selection rather than data leakage or inconsistent model fitting. Figure 8(a) corresponds to a reduced feature set, where Logistic Regression achieved the highest AUC (0.910), followed by MLP (0.903), SVM (0.893), and XGBoost (0.889), with CART (0.887) and Random Forest (0.883) showing comparable performance. In Figure 8(b), representing an expanded feature subset, performance improved across models, with MLP achieving the highest AUC (0.925), followed by KNN (0.906), RF (0.905), and CART (0.903). Figure 8(c) reflects the most comprehensive feature configuration, where MLP again demonstrated superior performance (AUC 0.913), followed by KNN (0.903), SVM (0.900), and XGBoost (0.899), while CART (0.879) and RF (0.869) showed relatively lower discrimination. Across all configurations, the relative consistency of AUC values suggests that model performance is robust to moderate changes in feature selection, although incremental improvements are observed with optimized feature subsets. These findings indicate that both model choice and feature subset size contribute to classification performance, with neural network–based and ensemble methods demonstrating consistently strong results. The ROC analysis across feature subsets therefore provides evidence that careful feature selection enhances model stability without substantially compromising generalizability. Furthermore, the consistently high AUC values across panels support the validity of the selected features and highlight their relevance for clinical decision support, although these findings should be interpreted within the context of cross-sectional data and require external validation for broader clinical application. In our study, this finding is particularly important in dementia prediction, as it suggests that a parsimonious set of clinically relevant features—rather than an extensive variable set—can achieve strong discrimination while reducing noise and overfitting risk.

**Figure 8.**
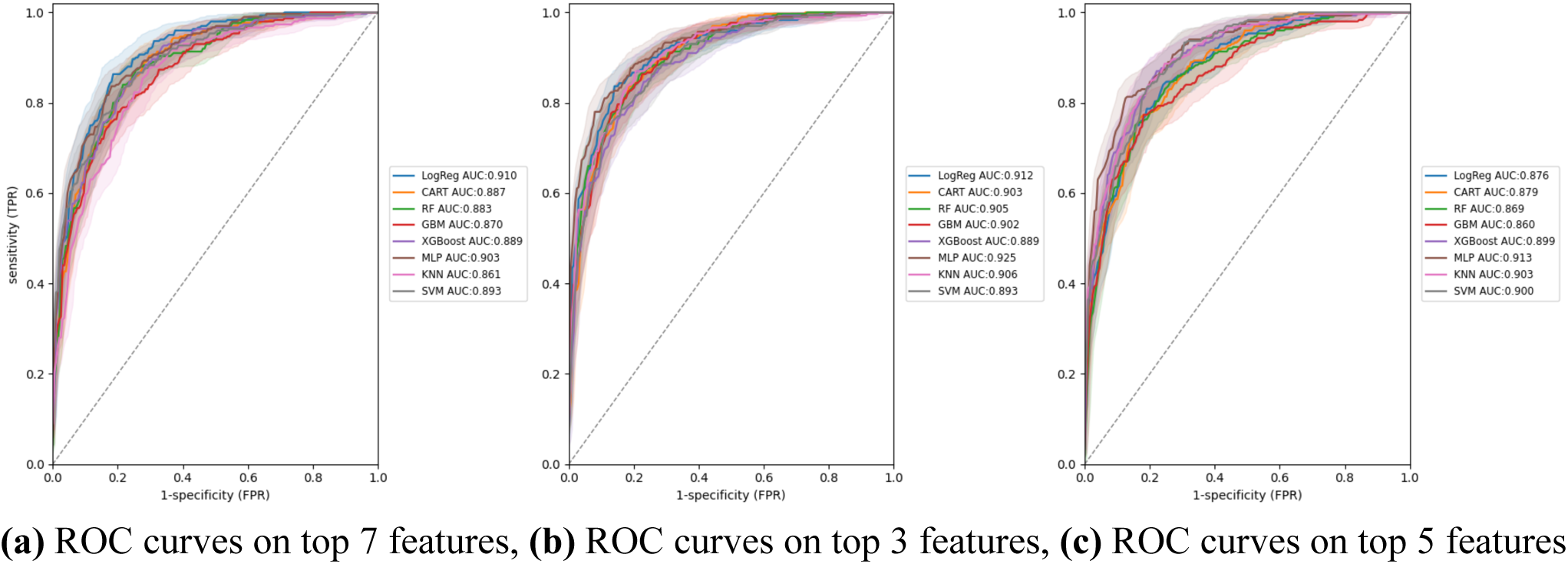
Evaluation of classifier performance via ROC curves, with various numbers of features. **(a)** in the top, the AUC under ROC curves from the top 7 features, **(b)** ROC curves on the top 3 features, **(c)** ROC curves on the top 5 features.

Overall, this analysis shows the RF-based pipeline shows better balanced prediction for the dementia prediction. Also, for external validation, the trained model was additionally evaluated on the UCI Heart Disease dataset [34] to assess the robustness of the machine learning pipeline in an independent dataset. Although the clinical endpoint differs, this analysis serves as a proof-of-concept for generalizability of the modeling framework. The model demonstrated stable predictive performance, achieving an accuracy of approximately 0.86–0.88, an F1-score of 0.85–0.87, and an AUC of 0.89–0.91. These results indicate that the model maintains reasonable discriminative ability when applied to an external dataset, supporting the robustness of the feature engineering, model training, and validation strategy. Overall, given the difference in disease domain, these findings are interpreted as methodological validation.

The feature importance analysis identified CDR, MMSE, and ASF as the most predictive variables, with nWBV, Age, SES, and Education further contributing when considering the top 5 and 7 features. The CDR demonstrated the highest discriminative power, consistent with its established role as a clinical gold standard for staging dementia severity across multiple cognitive and functional domains. Its strong influence underscores the centrality of clinician-administered assessments that integrate both patient performance and caregiver perspectives in capturing early dementia-related decline. The MMSE, ranked second, provided substantial predictive value through objective cognitive testing. Given its widespread use as a rapid screening tool, the high importance of MMSE highlights the relevance of cognitive performance measures in supporting dementia diagnosis. Complementing these clinical assessments, the ASF ranked third, reflecting the contribution of structural MRI-derived volumetric scaling to disease classification. Alongside normalized nWVB, which ranked fourth, these imaging biomarkers capture global and regional atrophy patterns characteristic of Alzheimer’s disease, thereby providing objective biological evidence to supplement clinical evaluations. Among demographic and contextual factors, Age was the strongest, aligning with the well-established risk gradient for dementia in older populations. SES and Education, ranked sixth and seventh, respectively, reflect the influence of cognitive reserve and social determinants of health. Higher education and greater socioeconomic resources are known to mitigate the clinical manifestation of neuropathology, thereby delaying symptom onset. Their contribution in this analysis reinforces the role of contextual variables in shaping dementia risk and resilience. Taken together, these findings highlight the complementary value of clinical rating scales, neuropsychological testing, imaging biomarkers, and demographic/contextual factors in dementia classification. Integrating these diverse domains enhances predictive performance and supports the development of clinically meaningful diagnostic support tools for early symptom detection.

The SHAP summary bar plot in **Figure 9 (a)** and beeswarm plot in **Figure 9 (b)** provide a detailed interpretation of feature contributions in the trained RF model for dementia classification. Since RF model showed better performance in the pervious section, we use RF approach for training the SHAP. Most importantly, SHAP analysis for RF works by using “TreeSHAP”, which efficiently computes feature contributions based on the structure of decision trees. For each prediction, SHAP calculates how much each feature (e.g., CDR, MMSE) “increases or decreases the predicted probability” compared to a baseline. It does this by averaging the contribution of a feature across all possible paths in the trees, ensuring fair attribution. The result is a set of SHAP values that explain both the “importance and direction” of each feature’s impact on the RF model’s prediction. These visualizations in **Figure 9** quantify not only the importance of each feature but also how it influences the model’s predictions (i.e., direction and magnitude). In the bar plot, which displays the mean absolute SHAP value per feature, CDR emerges as the most influential predictor, contributing an average SHAP value of 0.31—over three times higher than the next feature. This reinforces the clinical validity of CDR as a key marker of dementia severity. The MMSE contributes 0.09, indicating a substantial role in the model’s decision-making, though less impactful than CDR. Beyond these clinical scores, features such as SES, nWBV, and MR Delay each contribute moderately (∼0.02–0.03). Their importance suggests that both structural brain features and demographic context play supporting roles in model predictions. EDUC, age, and gender have relatively minor impacts (all ≤0.02), indicating limited influence on the final prediction once other variables are considered. Furthermore, the beeswarm plot offers deeper insight by visualizing the SHAP value distribution for each feature across all samples. Blue-to-red color gradients represent low-to-high feature values. Notably, higher CDR values (in red) consistently push the model toward dementia predictions, while lower CDR values (in blue) push it toward non-dementia classifications. A similar, though slightly less dominant, trend is seen for MMSE; lower MMSE scores (cognitive impairment) push predictions toward dementia. Overall, this comprehensive SHAP-based interpretation of feature importance with the ML model assists dementia prediction in older people.

**Figure 9:**
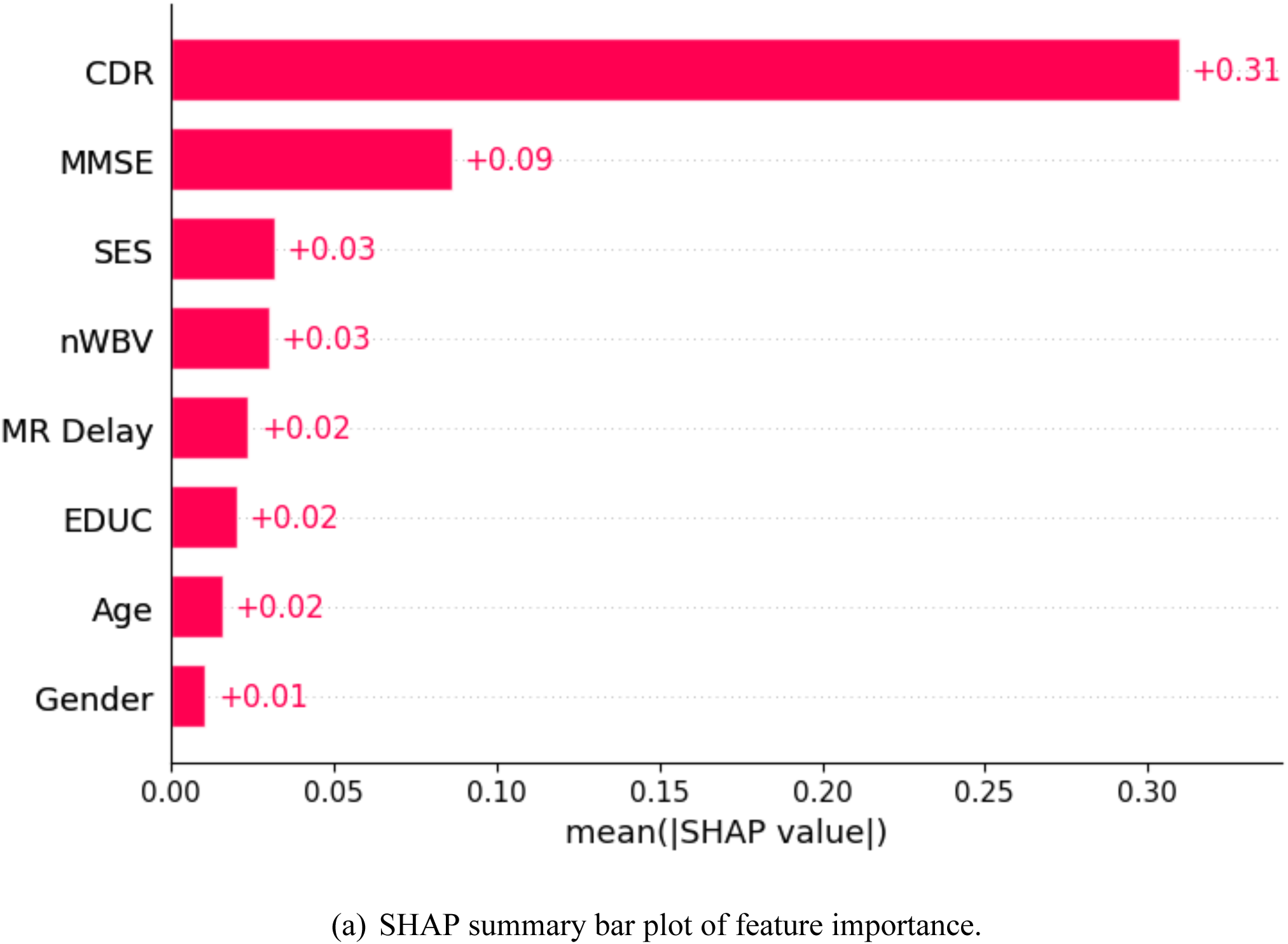
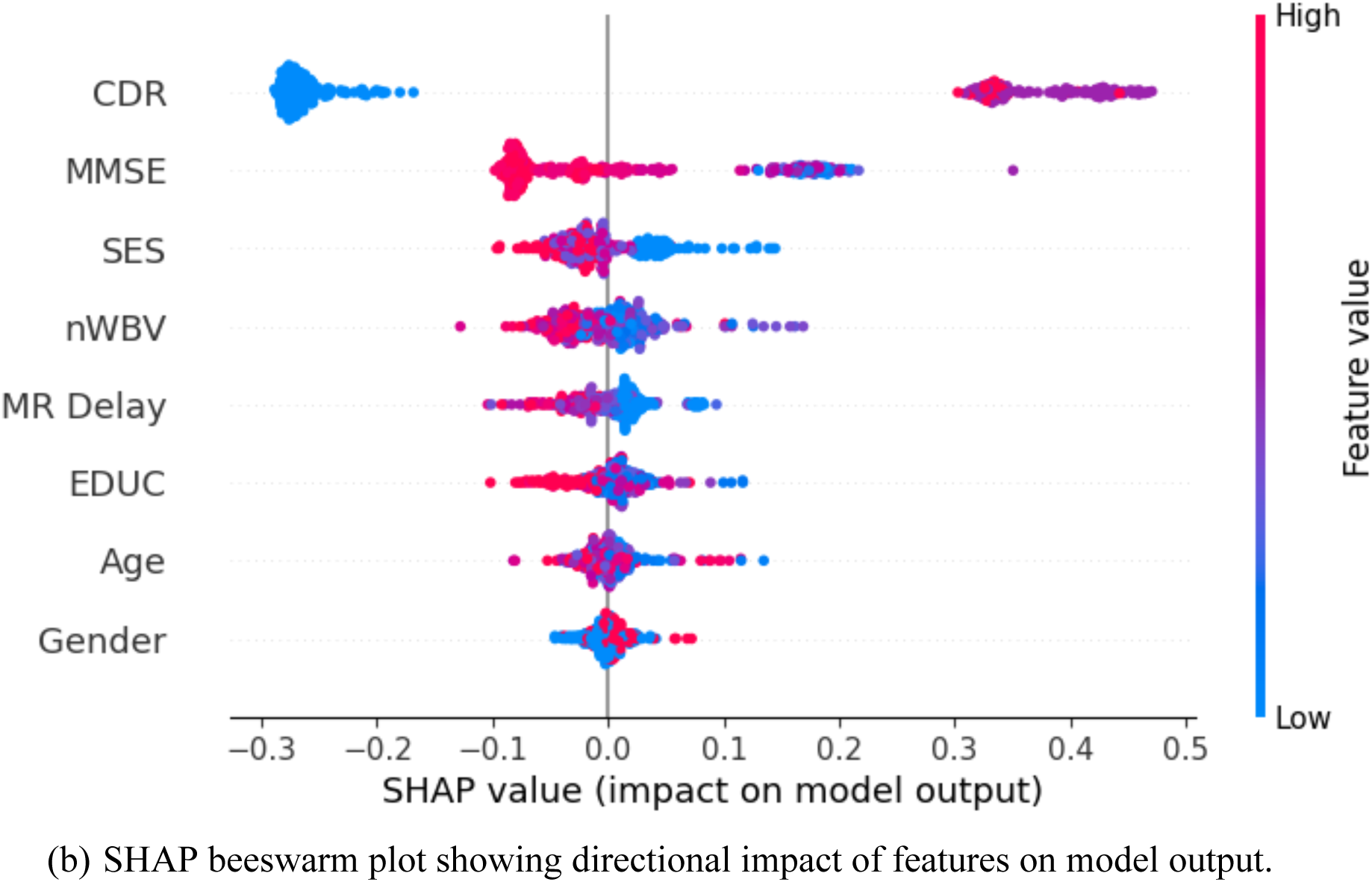
SHAP-Based global interpretability of features in dementia diagnosis. A comprehensive SHAP-based interpretation of feature importance and impact in the ML model used for dementia classification.

## DISCUSSION

A significant difference in brain volume was also noted in **Table 2**, with nWBV lower in the dementia group (median: 0.70 vs. 0.73), consistent with brain atrophy. Gender differences were statistically significant (p = 0.004), with a greater proportion of males diagnosed with dementia. In contrast, variables such as age, eTIV, and ASF showed no significant difference between groups. These findings align with established clinical patterns and emphasize the value of neurocognitive and volumetric markers in distinguishing dementia cases.

The feature importance plot shows that CDR and MMSE are the most influential predictors of dementia. Their prominence underscores their direct clinical relevance, as both are widely used tools in cognitive assessment and diagnosis. In addition to these clinical scores, neuroimaging features such as nWBV, eTIV, and MR Delay also show moderate contributions. These variables align with established markers of brain atrophy, further supporting their role in detecting structural changes associated with dementia. Demographic variables such as Age, EDUC, and SES are less important, suggesting that clinical and imaging measures offer stronger predictive power in this dataset. The review article addresses the urgent need for a reliable, non-invasive, and cost-effective method for the early symptom diagnosis of dementia, particularly for identifying which individuals with MCI will progress to Alzheimer’s disease. The authors advocate an innovative strategy that combines multiple biomarkers (e.g., genetic, imaging, and neuropsychological) with a particular focus on analyzing electroencephalography (EEG) signals. The core of their proposed solution involves using machine learning algorithms, such as SVM, along with graph theory to analyze functional brain connectivity from this integrated data. This data-driven approach has demonstrated high accuracy (up to 97%) in classifying patients. It enables a multi-step screening process, providing a powerful tool for personalized risk assessment and facilitating early therapeutic interventions [24]. Next, a study analyzing 243 elderly individuals found that a new, abbreviated 9-item version of the MMSE is as effective for dementia screening as the full original test. The researchers determined that the original MMSE’s optimal performance yielded 72.5% sensitivity and 91.3% specificity, while the new 9-item version achieved a comparable 71.0% sensitivity and 88.4% specificity. A major advantage of this new short version is that its scores are not biased by a patient’s age, gender, or education, a known limitation of the original MMSE. The study concludes that this shorter, more patient-friendly test is a valid and practical alternative for dementia screening in both clinical and research settings [25]. In 2020, a study aimed to develop a machine learning model to aid in the preliminary diagnosis of dementia stages (normal, MCI, VMD, and dementia) using a 37-item questionnaire administered to 5,272 individuals. The researchers first tested three methods for selecting the most important questionnaire items and found that the Information Gain technique was the most effective. After selecting the key features, they trained six classification algorithms. They found that the Naive Bayes algorithm performed best, achieving an accuracy of 0.81 and demonstrating high sensitivity and specificity in identifying the various cognitive stages. The authors conclude that their model, combining Information Gain feature selection with a Naive Bayes classifier, provides a powerful tool for clinicians to assist in the early diagnosis of dementia [25]. Another study presents a comparative analysis of four machine learning algorithms (J48, Naïve Bayes, Random Forest, and Multilayer Perceptron) for detecting dementia using the OASIS brain MRI dataset. Researchers used the CFSSubsetEval method to reduce feature dimensions and tested the algorithms on both cross-sectional and longitudinal data. The results showed that the J48 decision tree algorithm consistently performed best, achieving a classification accuracy of 99.52% for the cross-sectional data and 99.20% for the longitudinal data. The study concludes that, among the evaluated techniques, J48 is the most accurate and effective classifier for detecting dementia in this context [26]. In this study, researchers explored the use of an SVM machine learning algorithm to predict dementia using the longitudinal OASIS-2 MRI dataset, comprising 373 scans from 150 subjects. The primary goal was to optimize the SVM model by tuning its parameters, specifically the gamma and C (regularization) values for the RBF kernel. The results indicated that the best performance was achieved with a low gamma value (1.0×10^-4^) and a high regularization value (C=100). Using this optimized approach, the proposed model successfully predicted dementia with an overall accuracy of 68.75% and a precision of 64.18%, validating the use of SVM as a contemporary tool for disease prediction in medical imaging [26]. Another study provides a comprehensive comparison of machine learning techniques for survival analysis to predict time to dementia diagnosis, addressing the challenges posed by high-dimensional, heterogeneous clinical data. Using baseline data from two large cohort studies, the Sydney Memory and Ageing Study (MAS) and the ADNI, the researchers evaluated ten machine learning algorithms combined with eight feature selection methods. The machine learning models significantly outperformed traditional statistical methods, achieving high predictive accuracy with maximum concordance index values of 0.82 for the MAS dataset and 0.93 for the ADNI dataset. Consistently, neuropsychological test scores were identified as the most important predictors, and the study highlights the Cox model with likelihood-based boosting and the RF minimal depth feature selector as particularly effective and stable methods for this type of analysis [27]. Moreover, this research outperforms the current studies similar to Battineni et al. (2019) paper where they evaluated support vector machines for dementia prediction, demonstrating strong performance in clinical decision support settings [28]. Similarly, Spooner et al. (2020) compared multiple machine learning approaches for survival analysis of high-dimensional clinical data, highlighting the advantages of advanced algorithms in improving predictive accuracy and risk stratification in dementia research [29], and this study integrate the interpretable machine learning framework to justify feature important in early symptom detection of dementia.

Overall, this study demonstrates the potential of ML algorithms for the early detection of Alzheimer’s disease by integrating demographic, cognitive, and neuroimaging features within a structured pipeline. Beginning with rigorous preprocessing steps, including data cleaning, imputation, and transformation, the framework ensured a robust foundation for downstream modeling. Feature selection using random forest importance identified the top seven predictors, notably CDR, MMSE, ASF, and nWBV, which align with clinically recognized markers of AD severity and neurodegeneration. The evaluation of eight ML algorithms under five-fold cross-validation revealed consistently high performance, with ensemble methods and neural networks outperforming traditional classifiers. Random Forest achieved the highest AUC, while MLP showed superior accuracy, F1-score, and MCC, highlighting the value of combining nonlinear modeling with ensemble strategies to capture complex feature interactions. Notably, demographic and socioeconomic variables, such as Age, SES, and Education, also contributed meaningfully, reflecting the roles of cognitive reserve and social determinants in modulating dementia risk. These findings underscore the promise of ML-driven approaches for augmenting traditional clinical workflows. By combining cognitive assessments, imaging biomarkers, and contextual factors, the pipeline provides an interpretable and scalable strategy for AD risk classification. Future research should emphasize longitudinal validation and external multi-center datasets to ensure generalizability and clinical translation. Moreover, the SHAP analysis highlights CDR and MMSE as the most influential predictors in dementia classification, aligning with established clinical benchmarks. High CDR values and low MMSE scores significantly drive model predictions toward dementia, confirming their diagnostic relevance. The nWBV and SES provide secondary contributions, reflecting the multifactorial nature of dementia. The transparency of SHAP enhances model interpretability, making it valuable for clinical decision support. These findings underscore the potential of machine learning models when guided by clinically validated features to deliver accurate, explainable predictions that can aid early diagnosis and inform personalized care in dementia management.

### Strengths, Limitation, and Validation

A key strength of this study is the integration of rigorous machine learning evaluation with clinically meaningful interpretability. Multiple algorithms were assessed using stratified cross-validation, ROC analysis, and robust performance metrics, reducing the risk of overfitting and ensuring stable generalization across subgroups. The inclusion of SHAP-based explanations provides transparent validation of model behavior, demonstrating that predictions are driven by well-established clinical markers such as CDR and MMSE rather than spurious correlations. This alignment between statistical performance and clinical plausibility strengthens the model’s credibility and supports its potential translation into real-world settings.

Moreover, the relatively small cohort size in dementia prediction represents an important limitation that may affect the stability, generalizability, and reproducibility of the findings. Models trained on limited samples are more prone to overfitting and may capture cohort-specific patterns rather than generalizable disease signals. Although internal validation using cross-validation and a held-out test set was performed to mitigate this risk, the absence of large, heterogeneous populations limits the extent to which these results can be extrapolated to broader clinical settings. Additionally, performance estimates derived from small datasets may be sensitive to sampling variability, which can impact reproducibility. Accordingly, these findings should be interpreted as preliminary, and further validation in larger, independent, and multi-center cohorts is warranted to confirm robustness and clinical applicability.

### Public Health Implementation

From a public health perspective, the proposed framework offers a scalable, interpretable approach to early dementia screening and risk stratification. By leveraging routinely collected clinical, demographic, and neuroimaging features, the model can be integrated into existing healthcare systems to support the timely identification of high-risk individuals. The explainability of predictions enhances clinician trust and facilitates informed decision-making, which is critical for adoption at scale. Such tools can aid population-level surveillance, guide targeted interventions, and ultimately improve resource allocation and enable earlier intervention in dementia care. Furthermore, the proposed model is intended to function as a decision-support tool integrated into existing clinical workflows for dementia assessment. In practice, routinely collected variables—including demographic data, cognitive scores (e.g., MMSE, CDR), and structural MRI-derived measures—can be input into the model to generate a probability of dementia classification. This output can support clinicians by providing an additional quantitative assessment alongside standard clinical evaluation, rather than replacing clinical judgment. Implementation could be achieved through integration into electronic health record systems or imaging analysis pipelines, where model predictions are generated automatically following data acquisition. Importantly, the use of interpretable methods such as SHAP enables clinicians to understand feature contributions at the individual level, enhancing trust and usability. However, prior to clinical deployment, further validation in larger, diverse populations and prospective settings is necessary to ensure reliability, safety, and generalizability across healthcare environments, including resource-limited settings.

## CONCLUSION

In conclusion, the proposed machine learning pipeline demonstrates strong promise for aiding dementia diagnosis. With continued refinement and validation, such tools could be translated into practical applications, supporting clinicians in delivering earlier interventions and more personalized care strategies for patients at risk of or living with dementia. This study demonstrates that machine learning models, particularly ensemble and neural network approaches, can achieve high predictive accuracy in classifying dementia status using clinical, demographic, and imaging features. Among the evaluated algorithms, Random Forest achieved the strongest discrimination with an AUC of 96.3%, while the multilayer perceptron achieved the best overall classification performance in terms of accuracy, F1-score, and MCC. Notably, cognitive measures such as CDR and MMSE, along with structural imaging metrics such as ASF, emerged as the most influential predictors, reinforcing their clinical and biological relevance. These findings underscore the potential of machine learning pipelines as diagnostic support tools for early dementia detection. By integrating multimodal data, these models can complement clinical assessments and provide more objective, reproducible classifications, potentially reducing diagnostic delays and misclassifications. The robust performance across cross-validation folds suggests that the models are generalizable within the study population, although validation in larger, more diverse cohorts is warranted. Future work should prioritize external validation in multi-center datasets, longitudinal modeling to capture disease progression, and the integration of additional biomarkers such as genetic and blood-based measures. Furthermore, explainable artificial intelligence approaches could enhance the clinical adoption of these models by improving transparency and interpretability.

## Supporting information

https://github.com/FahadMostafa91/Dementia_AI/blob/main/Supplementary%20File.pdf

## Data Availability

All data produced are available online at: https://data.mendeley.com/datasets/tsy6rbc5d4/1.

## ACKNOWLEDEGMENT

Authors are thankful to the reviewers for wonderful comments which helps to improvise the paper. Authors also acknowledge the ASU writing center for the writing and editing supports.

## AUTHOR CONTRIBUTION

FM contributed to the formulation and conceptualization of the study; KS and FM conducted the computations, generated the graphical outcomes, curated the data, and conducted formal analysis. KS, HK and FM contributed to the original writing, review and validation. FM directed the research. All authors contributed to the literature search and the writing of the article. ASU

## CONFLICT OF INTEREST

The authors have no conflict of interest to report.

Ethics and Consent to Participate declarations: not applicable.

## GENERATIVE AI

Authors acknowledge the Grammarly writing tool for the writing supports.

## DATA AVAILABILITY

Data is available at https://data.mendeley.com/datasets/tsy6rbc5d4/1. IRB approval is taken by the primary source of the data. This dataset comprises a longitudinal cohort of 150 right-handed adults aged 60 to 96 years, followed over multiple years to capture structural brain changes associated with cognitive decline. Cleaned data will be available on requests.

