## Supplementary material for "Development and Validation of Dementia Diagnosis in Adults through Machine Learning Frameworks: A Cross-Sectional Study Using Clinical and Imaging Data": https://github.com/FahadMostafa91/Dementia_AI/blob/main/Supplementary%20File.pdf

### Supplementary File

This **Table 7** summarizes the performance of eight machine learning models evaluated under three feature scenarios: (1) using only CDR and MMSE, (2) excluding CDR, and (3) excluding MMSE. Performance metrics include Accuracy, Kappa, Sensitivity, Precision, F1 Score, and AUC for both cross-validation (CV) and independent test sets. Results show that models using both CDR and MMSE achieve near-perfect performance. Excluding CDR leads to a substantial decline in performance, highlighting its importance as a key predictor. In contrast, excluding MMSE has a relatively minor effect, suggesting that CDR contributes more strongly to the model's predictive power.

**Table 7**: Performance of ML models evaluated under three feature scenarios.

| Scenario | Model | CV Acc | CV Kappa | CV Sens | CV Prec | CV F1 | CV AUC | Test Acc | Test Kappa | Test Sens | Test Prec | Test F1 | Test AUC |
|---|---|---|---|---|---|---|---|---|---|---|---|---|---|
| CDR+MMSE | RF | 0.99 | 0.98 | 0.98 | 1.00 | 0.99 | 1.00 | 1.00 | 1.00 | 1.00 | 1.00 | 1.00 | 1.00 |
| CDR+MMSE | SVM | 0.99 | 0.98 | 0.98 | 1.00 | 0.99 | 1.00 | 1.00 | 1.00 | 1.00 | 1.00 | 1.00 | 1.00 |
| Exclude CDR | RF | 0.74 | 0.48 | 0.71 | 0.77 | 0.74 | 0.82 | 0.93 | 0.87 | 1.00 | 0.88 | 0.94 | 0.97 |
| Exclude CDR | SVM | 0.81 | 0.62 | 0.69 | 0.92 | 0.79 | 0.86 | 0.73 | 0.47 | 0.67 | 0.77 | 0.71 | 0.83 |
| Exclude MMSE | RF | 0.99 | 0.98 | 0.98 | 1.00 | 0.99 | 1.00 | 1.00 | 1.00 | 1.00 | 1.00 | 1.00 | 1.00 |
| Exclude MMSE | SVM | 0.98 | 0.97 | 0.98 | 0.98 | 0.98 | 0.99 | 1.00 | 1.00 | 1.00 | 1.00 | 1.00 | 1.00 |
